# Low-branching vessel architecture shapes immune cell niches and predicts immune responses in renal cancer

**DOI:** 10.1101/2025.06.05.25329017

**Authors:** Marieta I. Toma, Yangping Li, Melina Kehl, Tim N. Kempchen, Laura Esser, Katharina Baschun, Sonia Leonardelli, Thomas Pinetz, Roberta Turiello, Michelle C.R. Yong, Natalie Pelusi, Guillermo Altamirano-Escobedo, Eduardo Bayro-Corrochano, Sebastian Kadzik, Markus Eckstein, Lukas Flatz, Fabian Hörst, Jens Kleesiek, Jonas Saal, Glen Kristiansen, Manuel Ritter, Jörg Ellinger, Viktor Grünwald, Niklas Klümper, Alexander Effland, Michael Hölzel

**Author notes:** these authors contributed equally (Marieta I. Toma, Alexander Effland, Michael Hölzel).

## Abstract

Clear cell renal cell carcinoma (ccRCC) is characterized by marked histological heterogeneity, encompassing its vasculature. Here, we introduce PropSegNet, a learning-based algorithm that segments and classifies three distinct vascular patterns in CD31-stained tissue sections. Integrating transcriptomic features, our work identifies a trajectory from high- to low-branching vessel architecture that co-evolves with the loss of proximal tubular cell traits in tumor cells. Furthermore, low-branching vessels form niches enriched with T cells and antigen-presenting cells. Mechanistically, air-liquid-interface cultures of patient-derived tumor fragments confirm that low-branching vessel features associate with T cell infiltration resulting in reduced viability under IL-2 rich conditions. Post-hoc transcriptomic analyses from two phase III clinical trials demonstrate that patients with tumors exhibiting an inflamed, low-branching vascular phenotype benefit most from the addition of immune checkpoint inhibition to anti-angiogenic treatment. These findings provide a rationale for prospective evaluation of vascular patterns and vessel-immune cell niches as potential biomarkers in ccRCC.

## Introduction

Clear cell renal cell carcinoma (ccRCC) is the most common subtype of renal cancer, comprising for approximately 70-80% of all kidney cancers. ccRCC is characterized by distinct histological features, like clear cell cytoplasm, resulting from the accumulation of lipids and glycogen, dissolved during tissue processing^1^. ccRCC is morphologically heterogeneous and the high phenotypic variability was linked to prognosis and therapy response^2^. Verine et al. proposed a new grading system, based on the architectural features^3^. Subclonal loss of both driver genes in ccRCC BAP1 and PBRM1 is related to particular tumor morphology, like rhabdoid features^4^. ccRCC shows also a remarkable intratumoral genetic heterogeneity^5^ and based on the clonal evolution, seven evolutionary subtypes of ccRCC, with distinct clinical outcome were described^6^.

The inactivation of the VHL (von Hippel-Lindau) tumor suppressor in up to 90% of cases of ccRCC leads to the dysregulation of the hypoxia-inducible factor (HIF) pathway^7^, promoting angiogenesis and tumor growth through the upregulation of vascular endothelial growth factor (VEGF)^8^. ccRCC frequently exhibit delicate vascular networks permeating the tumor tissue. The microvessel density in ccRCC was subject to numerous studies and has been proposed to be predictor for good prognosis^9,10^. Other studies have examined the heterogeneity of the vessel architecture in renal cancers, proposing partially overlapping classifications^11–13^. However, these classifications have not been linked to distinct molecular characteristics or clinical outcomes.

Overall, ccRCC is a heterogeneous disease, with prognosis and treatment responses influenced by both histopathological and molecular factors, emphasizing the need for personalized treatment approaches. The current treatment landscape of advanced stage ccRCC centers on immune checkpoints inhibitors (ICI) and anti-angiogenic drugs blocking VEGF signaling, and combinations thereof^14,15^. Sequencing of treatments^16^ and the identification of predictive biomarkers for better patient stratification remain a pivotal focus of ongoing research^17–19^. Biomarkers would be of significant clinical value to identify metastatic ccRCC (mRCC) patients who are unlikely to benefit from the addition of ICI, which carries a risk of life-threatening toxicity, to anti-VEGFR tyrosine kinase inhibitors (TKI). This is particularly relevant given that the benefit of incorporating ICI for improving long-term survival remains questionable especially in angiogenic-high mRCC, which are overrepresented in the IMDC (International Metastatic RCC Database Consortium) low-risk group^17,19,20^. PD-L1 receptor status, routinely determined immunohistochemically in therapy-naive tumor tissue, plays a minor role in mRCC therapy selection, as its predictive performance is inconclusive^21^. Furthermore, therapeutic benefit of ICI+TKI combination therapy was observed regardless of PD-L1 status^22^, and thus current guidelines do not recommend PD-L1 status for mRCC treatment decisions.

Here, we developed a novel learning-based algorithm for the classification of vascular patterns in ccRCC, which allows for a quantification of respective areas. Assigning transcriptional signatures to vascular patterns revealed co-evolution with tumor cell differentiation and differences in the therapeutic benefit from ICI therapy. Moreover, air-liquid interface (ALI) cultures of patient-derived tumor fragments (PDTF) recapitulated these associations between vascular patterns and tumor-intrinsic immune phenotypes. Finally, multiplexed immunofluorescence imaging supported a prominent role of vessel-immune cell niches for tumor-intrinsic immunity linked to a low-branching vessel architecture in ccRCC.

## Results

### Automated segmentation and classification of vascular patterns

We used immunohistochemistry (IHC) to detect endothelial cells by CD31 expression in ccRCC tissue sections and we determined three distinct vascular patterns as high-branching (HB), low-branching (LB), and sinusoidal (SN) (**Fig. 1a-c**). The HB pattern was characterized by a highly branched network of small vessels surrounding small nests of tumor cells, whereas the LB pattern exhibited elongated vessels within larger groups or sheets of tumor cells. Large dilated vessels were characteristic for the SN pattern. Since we observed co-occurrence of the three vascular patterns within tumors, we devised an unbiased, automated approach. For the automatized segmentation and subsequent classification of the three vascular patterns, we propose a novel learning-based algorithm termed Proportion Segmentation Network (PropSegNet). As visualized in **Fig. 1d**, CD31 IHC image data with the usual RGB color scheme are the input of the Swin Transformer-type neural network, which is a hierarchical transformer exhibiting a shifted window scheme. The network generates pixel-wise segmentation masks representing the individual pattern structures. The proportion of each pattern is subsequently calculated based on the relative size of these segmentation masks. We emphasize that only the global proportion of each pattern is available during training, i.e., the percentage of the pixels in the tissue covered by the specific pattern. A detailed description of PropSegNet, the adoption of existing methods^23–25^ and the training datasets is provided in Supplementary Methods and Supplementary Table 1.

**Figure 1.**
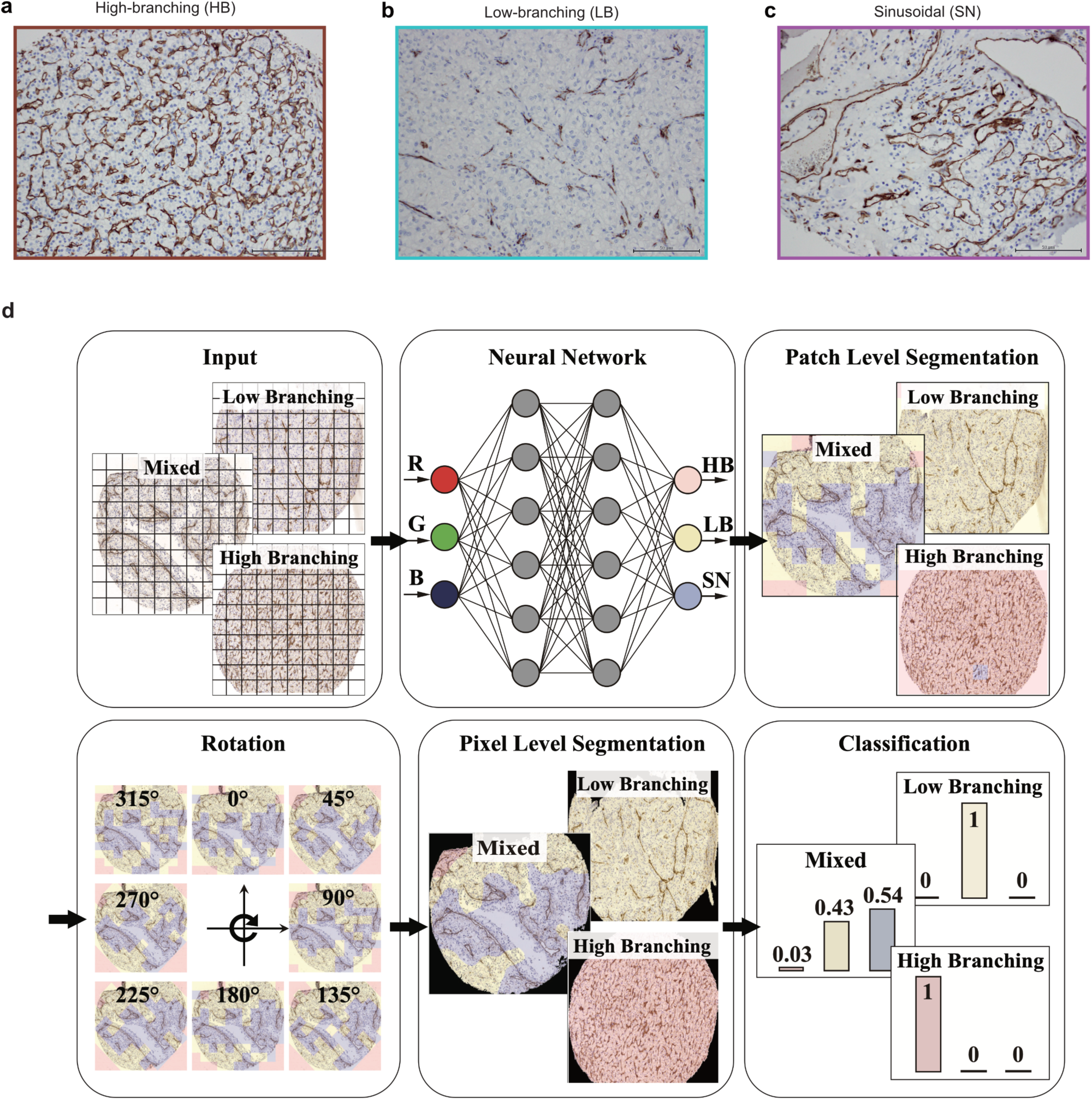
Automated segmentation and classification of distinct vascular patterns in ccRCC tissue sections by PropSegNet. Representative examples show sections with predominant **(a)** high-branching, **(b)** low-branching, and **(c)** sinusoidal vascular patterns in ccRCC, as identified by CD31 immunohistochemistry. Scale bars represent 50 µm. **(d)** Schematic representation of PropSegNet, a learning-based algorithm for automated segmentation and subsequent classification of vascular patterns.

### Co-evolution of vascular patterns and tumor cell lineage traits

We then applied PropSegNet to CD31 IHC image data from a cohort of advanced-stage ccRCC patients (67 samples from 61 patients: 48 primary tumors, 6 paired distant metastases, 13 unpaired distant metastases), and determined the proportions of the HB, LB and SN vascular patterns for each sample (**Fig. 2a**). Patient characteristics are detailed in Supplementary Table 2 and 3. Subsequently, we ranked the samples by decreasing proportion of the HB vascular pattern, uncovering a negative correlation between the HB and LB vascular patterns as the most significant relationship (**Fig. 2b**). To assign molecular features to the different vascular patterns, we isolated total RNA from matched formalin-fixed, paraffin-embedded (FFPE) ccRCC tissue samples and conducted 3’mRNA sequencing. We then correlated gene expression levels with the content of the vascular patterns and selected genes with the highest rank correlation values for compiling gene signatures associated with the HB, LB and SN patterns (**Fig. 2c** and Supplementary Table 4). As the number of correlating genes was highest for the HB vascular pattern, we applied a more stringent cutoff value to limit the number of genes as recommended for downstream analysis like gene set enrichment analysis (GSEA)^26^. We also confirmed that the gene signatures were consistently correlated with the output of PropSegNet (Supplementary Fig. 1).

**Figure 2.**
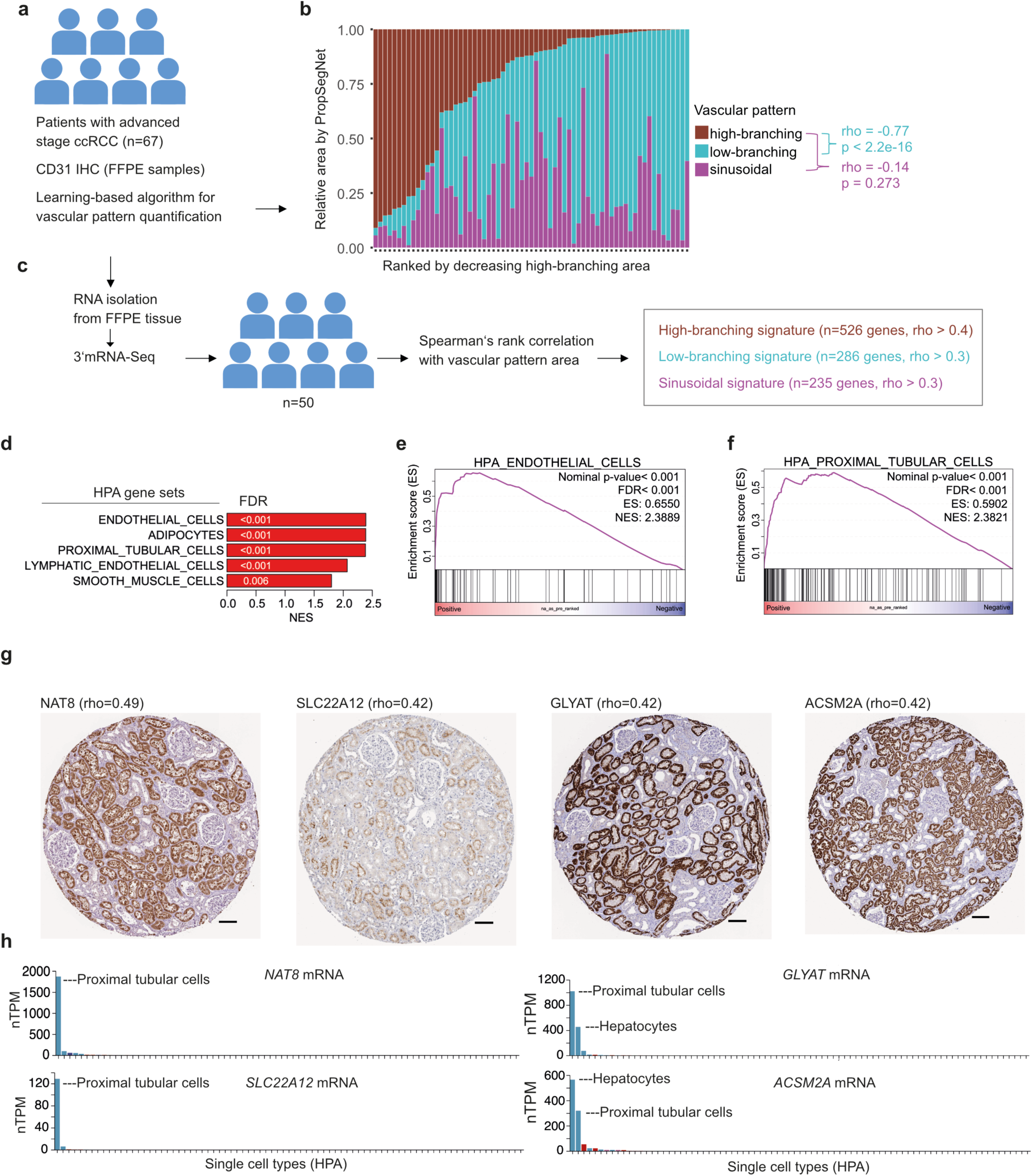
Co-evolution of vascular patterns and tumor cell lineage traits in ccRCC. **(a)** Overview of ccRCC tissue specimens used for analyses by PropSegNet. **(b)** Stacked barplot visualizing relative area of indicated vascular pattern per sample as quantified by PropSegNet. Spearman’s rank correlations (rho) between vascular patterns are indicated. **(c)** Summary of strategy to assign transcriptome features to vascular patterns using matched samples. **(d)** Top ranking gene sets from GSEA associated with the high-branching vascular pattern. Cell type genes signatures from the Human Protein Atlas (HPA) project were used as gene set collection for GSEA. Individual GSEA plots shown for **(e)** endothelial cells and **(f)** proximal tubular cells. **(g)** Normal kidney stained for indicated proteins encoded by core enrichment genes related to the GSEA plot for the proximal tubular cell gene signature shown in (f). Images were taken from the HPA website (https://www.proteinatlas.org/). Links to source images from the HPA project are provided in Supplementary Figure 2. Scale bars represent 100 µm. **(h)** Normalized expression of indicated core enrichment genes in the single cell type dataset from the HPA project. Data and visualization were downloaded from the HPA website and modified.

Next, we carried out gene set enrichment analysis (GSEA) using the gene correlation values as ranked metric input and the gene set collection of the Human Protein Atlas (HPA) project focused on cell types^27,28^ (Supplementary Table 5). As expected, we found that the top-ranking cell type gene sets associated with the HB vascular pattern were endothelial cells and smooth muscle cells, the latter reflecting likely abundant pericytes in ccRCC (**Fig. 2d, e**). Notably, the HB vascular pattern was also associated with the proximal tubular cell lineage (**Fig. 2d, f**). Applying leading-edge analysis to GSEA, we selected a few candidate genes (*NAT8, SLC22A12, GLYAT, ACSM2A*) and verified their selective protein and mRNA expression in proximal tubular cells in the public HPA database (**Fig. 2g, h** and Supplementary Fig. 2). Together, the results reveal an association between vascular patterns and tumor cell states. In particular, ccRCC with a HB vascular phenotype being characterized by high expression of proximal tubular cell genes, while ccRCC with a LB vascular phenotype showed downregulation of this cell lineage trait.

### Vascular pattern transcriptional signature predicts response to ICI therapy

To further validate our findings and explore clinical relevance, we analyzed two independent transcriptome datasets derived from the JAVELIN Renal 101 and IMmotion 151 phase III randomized trials, both of which evaluated the addition of ICI to VEGF signaling blockade^14,15,17,19,20^. The JAVELIN Renal 101 trial compared avelumab plus axitinib versus sunitinib, and the IMmotion151 trial compared atezolizumab plus bevacizumab versus sunitinib. Avelumab and atezolizumab are ICI that block PD-L1, whereas axitinib, sunitinib and bevacizumab target angiogenesis by inhibiting VEGF signaling. In both cohorts, the HB signature correlated with gene sets of endothelial and proximal tubular cells (**Fig. 3a-d**), confirming the results from our discovery cohort. We then investigated associations between expression of the HB-signature and treatment responses (progression free survival; PFS). To this end, we divided the cohorts by a median classifier into HB-signature UP (HB.UP) and HB-signature DOWN (HB.DOWN) subgroups, assuming for the latter a predominant LB phenotype. Within the subgroups, we then analyzed therapy responses (PFS) stratified by the different treatment arms of the trials (**Fig. 3e-h**). Notably, benefit from the addition of ICI to VEGF signaling inhibition was restricted to the HB.DOWN subgroups in both cohorts with hazard ratios (HR) of 0.45 (95% CI: 0.33-0.61, log-rank p=1.65e-07) and 0.64 (95% CI: 0.51-0.80; log-rank p=0.00013), respectively (**Fig. 3f, h**). In contrast, there was no apparent benefit from ICI for the HB.UP subgroups, HR 0.98 (95% CI: 0.71-1.35) and HR 1.01 (95% CI: 0.78-1.32) (**Fig. 3e, g**). Comparing the median PFS of the sunitinib arms between the subgroups, patients with HB.UP tumors showed a more favorable response (mPFS: 11.1 and 12.6 months) than patients with HB.DOWN tumors (mPFS: 5.6 and 5.7 months). Previously, Motzer and colleagues used non-negative matrix factorization (NMF) clustering to define molecular subsets and described seven NMF clusters that were associated with treatment responses in the IMmotion151 trial^17^. We ranked the IMmotion151 cohort by decreasing expression of the HB-signature and, as expected, we found that the angiogenic and angiogenic-stromal NMF clusters were strongly enriched in the HB.UP subgroup (**Fig. 3i**). The five other NMF clusters, previously assigned to proliferative and inflammatory phenotypes, were significantly associated with the HB.DOWN subgroup (**Fig. 3i**). Using additional transcriptional signatures described by Motzer and colleagues, we further confirmed that the HB.DOWN subgroup was associated with proliferation and inflammation, in particular myeloid cells and the complement pathway (**Fig. 3j**). In summary, the results suggested that ccRCC with a predominant LB vascular pattern phenotype (HB.DOWN subgroup) were more immune cell infiltrated (inflamed TME), which aligns with and may explain the enhanced response to ICI-containing regimens observed in this subgroup. However, our findings were only correlative, and therefore we decided to employ patient-derived 3D tumor models to provide additional functional evidence.

**Figure 3.**
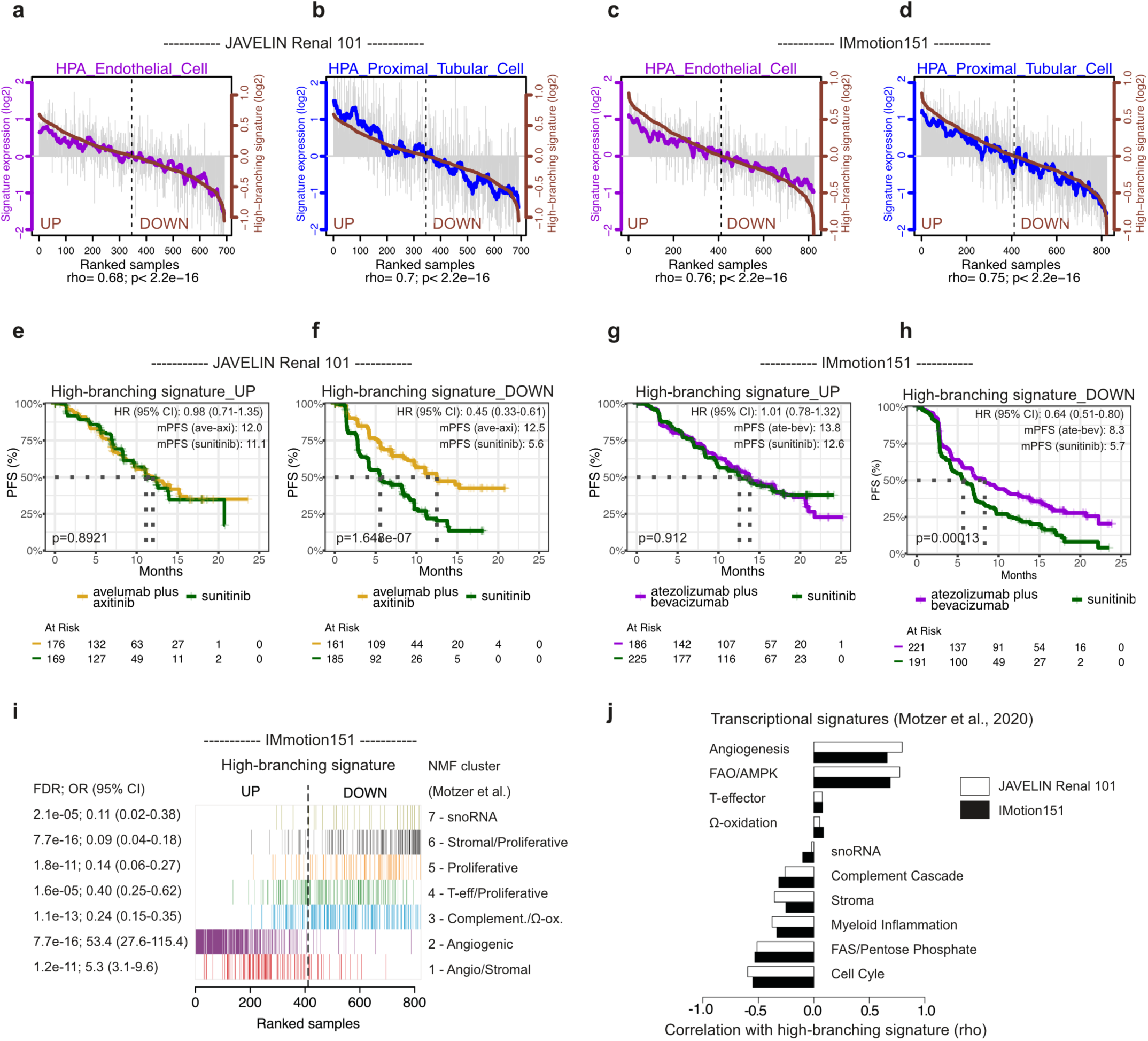
Patient stratification by vascular pattern gene signature predicts response to immune checkpoint inhibitor in post-hoc analyses of two clinical trials. **(a-b)** Correlation of the high-branching (HB) signature (brown) with the (a) HPA endothelial cell signature (violet) and the (b) HPA proximal tubular cell signature (blue) in the JAVELIN Renal 101 cohort. Samples ranked by decreasing expression of the HB-signature. Spearman’s rank correlation values (rho) as indicated. Vertical grey bars indicate expression of the endothelial or proximal tubular cell signature in individual samples. Violet or blue colored line indicates moving average to visualize trend. **(c-d)** Same analyses and visualization as described in a-b, but for the IMotion151 cohort. **(e-f)** Analyses of progression-free survival (PFS) in the JAVELIN Renal 101 cohort stratified first by median classifier for HB-signature expression, UP (e) vs. DOWN (f), and then by treatment arms avelumab plus axitinib versus sunitinib. Hazard ratios (HR) with confidence intervals (CI), p-value for log-rank test as indicated. **(g-h)** Same analyses as described for e-f, but for the IMotion151 cohort with the treatment arms atezolizumab plus bevacizumab versus sunitinib. **(i)** Association between expression of the HB-signature and the different NMF transcriptional clusters described by Motzer *et al.*^17^ visualized by barplots and samples of the IMotion151 cohort ranked by decreasing HB-signature expression. Two-sided Fisher’s exact test. FDR: false discovery rate, OR: odds ratio with 95% confidence interval. **(j)** Correlation (rho, Spearman’s rank correlation) of the HB-signature with additional transcriptional signatures described by Motzer et al.^17^ in the JAVELIN Renal 101 and IMotion151 cohorts.

### Vascular patterns associate with viability of PDTF cultures

Neal and colleagues described an air-liquid interface (ALI) method for improved culturing of patient-derived tumor fragments (PDTF) and modelling of the tumor microenvironment^29^. The published protocol involves adding IL-2 to the culture medium to maintain T cells within the tumor tissue samples, though this effect is limited to a few weeks. In a pilot study involving a limited number of cases, we previously confirmed that the ALI culture method is suitable for propagating patient-derived tumor fragments (PDTF) with stromal and immune cells, at least for some time^30^. In the present study, we established a cohort of 30 cases (Supplementary Table 6). ALI cultures were established from primary tumors after surgical removal according to the published protocol by Neal and colleagues, requiring 3 weeks on average. Once multiple characteristic round, organoid-like structures had formed, the ALI cultures were passaged and subsequently subjected to a one-week treatment with nivolumab, cabozantinib or left untreated (**Fig. 4a**). To assess viability and to determine potential treatment responses, we had explored different methods (e.g. ATP-based, flow cytometry), but finally decided for serial sectioning of the fixed ALI cultures (FFPE) followed by H&E staining and pathological scoring of the viable tissue area. By employing this approach, we were confident to account for the inherent variability in the input material, including confounding debris and capturing all available tissue fragments in an unbiased manner. Notably, we observed substantial variability in viability already of untreated ccRCC ALI cultures and **Fig. 4b** presents examples of H&E-stained tissue sections with high, intermediate and low viability, respectively. The quantification across all cases revealed viability ranging from 8% to 100%, with a median of 52.5% (**Fig. 4C**). Given that the considerable variability limited the assessment of treatment responses, we first aimed to identify correlates and elucidate the underlying mechanisms driving this heterogeneity. We found that ccRCC ALI cultures with high viability showed a higher proportion of the HB vascular pattern in the corresponding primary tumors as determined by PropSegNet (**Fig. 4d**). Conversely, ccRCC ALI cultures with low-viability showed a higher proportion of the LB and SN vascular patterns (**Fig. 4e, f**). This result was unexpected, as the LB vascular pattern phenotype was associated with a more aggressive clinical behavior and enhanced proliferation in the JAVELIN Renal 101 and IMmotion 151 cohorts (**Fig. 3i, j**). Of note, the ALI cohort also exhibited a higher proportion of cases with a predominant HB vascular pattern when compared to our advanced stage cohort (**Fig. 2a**), a difference that can be attributed to the clinical context, as patients undergoing surgical removal of primary ccRCC are more likely to have early-stage disease. We also conducted NGS amplicon sequencing for known ccRCC driver mutations, showing a significant association of BAP1 mutations with the LB vascular phenotype and a trend for low viability in ccRCC ALI cultures (**Fig. 4g** and Supplementary Table 7). In conclusion, the findings suggest that the variability in viability of ccRCC ALI cultures associated with vascular pattern phenotypes, which encouraged us to conduct a more comprehensive characterization.

**Figure 4.**
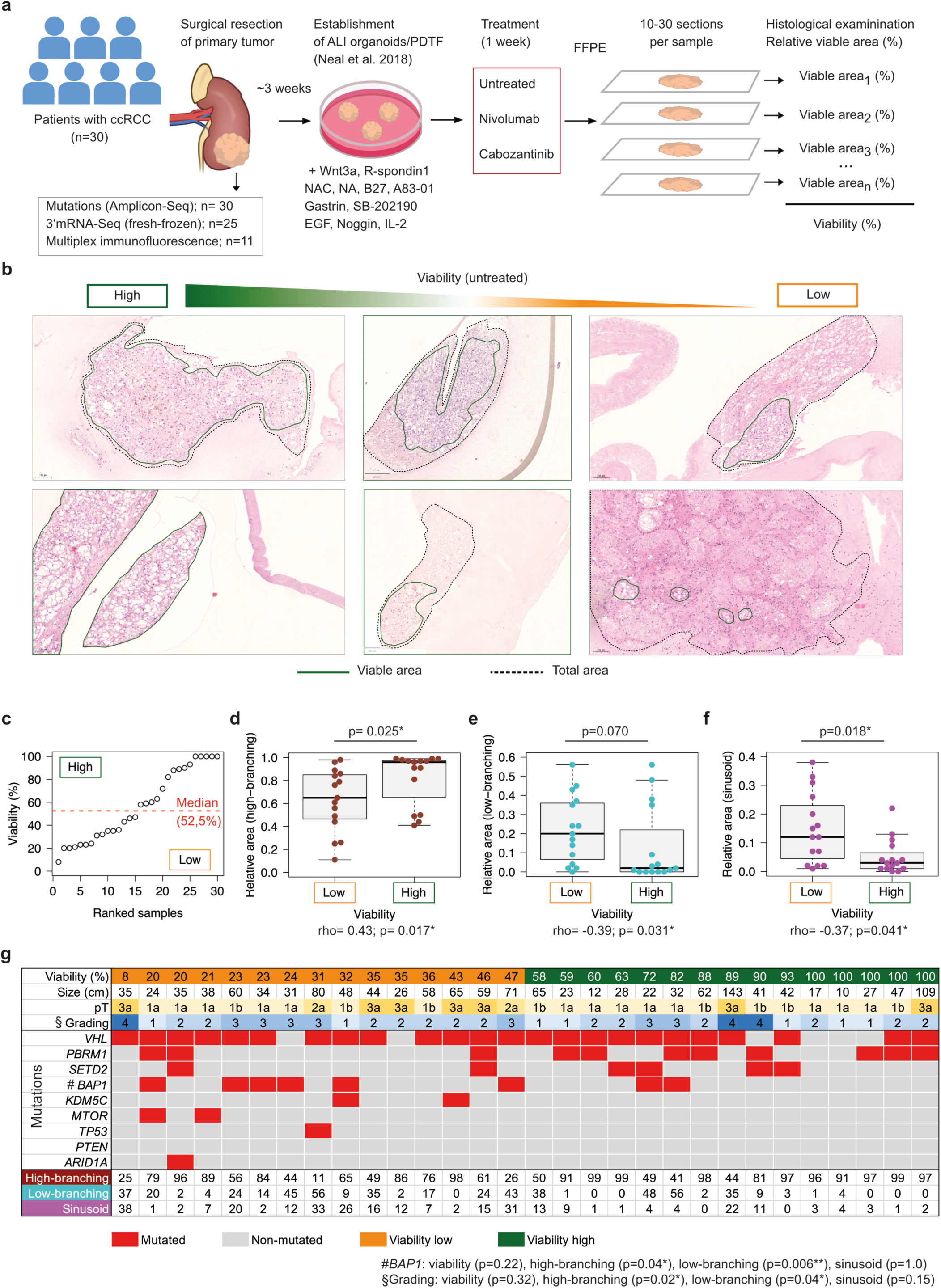
Association of vascular patterns with viability in air-liquid-interface cultures of ccRCC patient-derived tumor fragments. **(a)** Workflow of establishing air-liquid-interface (ALI) cultures of ccRCC patient-derived tumor fragments (organoids) following the protocol by Neal et al.^29^ and assessment of tumor tissue viability. **(b)** H&E stains showing representative examples of ALI cultured PDTF with high (left panels), intermediated (middle panels), and low viability (right panels). Scale bars represent 100 µm or 200 µm as indicated. **(c)** Summary of viability (% viable tumor tissue) across cohort (n=30) and unbiased median classifier for low versus high viability as indicated. **(d-f)** Association viability status with (d) high-branching, (e) low-branching, and (f) sinusoid vascular pattern of corresponding primary tumors. Group comparisons by two-sided Wilcoxon test, and Spearman’s rank correlation values for ungrouped comparisons. **(g)** Table summarizing key clinical parameters and ccRCC driver mutations detected in the cohort.

### T cell and chemokine transcript levels associate with reduced viability

From 25 out of 30 cases (83%) we were able to archive fresh-frozen tissue in parallel for RNA isolation and 3’mRNA sequencing. We compared the transcriptomes of the primary tumors corresponding to ALI cultures with low-(n=11) versus high-viability (n=14) and conducted GSEA using two widely used gene set collections (**Fig. 5a**). Analysis of the top enriched gene sets revealed that primary tumors of low-viability ALI cultures were characterized by enrichment of inflammatory and immune cell gene sets, as well as those related to cell cycle and proliferation (**Fig. 5b-c**). More specifically, the enriched genes sets were from T cells, B cells, plasma cells, macrophages, antigen presentation-related (allograft rejection) (**Fig. 5d**) and interferon responses (**Fig. 5e**), collectively indicating an inflamed TME. Conversely, primary tumors of high-viability ALI cultures were characterized by enrichment of angiogenesis, endothelial cell and fibroblast or pericyte gene sets, as well as those related to epithelial-mesenchymal transition (EMT), stress, TGF-beta, WNT and TNF signaling (**Fig. 5f-i**). At the individual transcript level, T cell, chemokine and cytokine transcripts were among the most differentially expressed genes, with higher levels in primary tumors of low-viability ALI cultures (**Fig. 5j**). Specifically, the higher expression of co-stimulatory T cell receptors such as *TNFRSF9* (CD137) and *CD226* (DNAM-1), along with Th1-type chemokines like *CXCL9* and cytotoxicity-related granzymes (*GZMA*, *GZMK*), suggested enhanced T cell-mediated anti-tumor immunity in low-viability ALI cultures (**Fig. 5k**). In contrast, we detected higher expression of immediate-early response genes of the AP-1 (activator protein-1) family (*FOS*, *FOSL1*, *FOSB*, and *JUNB*) and EGR (early growth response) family (*EGR1*, *EGR2*, and *EGR3*) in the primary tumors of high-viability ALI cultures (**Fig. 5j, l**). The AP-1 and EGR families of transcription factors are involved in regulating gene expression in response to external stimuli, such as growth factors or stress. Consistent with the predominant HB vascular pattern phenotype of high-viability ALI cultures (**Fig. 4d**), endothelial cell expressed transcripts (e.g. *ENG*, *EMCN*, *ESAM*) were also found at higher levels (**Fig. 5l**). In summary, our results demonstrated that elevated T cell and chemokine transcript levels were associated with reduced viability in corresponding ccRCC ALI cultures. This suggests that enhanced tumor-intrinsic immunity and T cell-mediated cytotoxicity, particularly under the IL-2-rich conditions employed in the standard ALI protocol, may contribute to lower culture viability observed in these ALI cultures. Moreover, as primary tumors of low-viability ALI cultures were enriched for the LB and SN vascular patterns, our findings are consistent with the re-analyses of the JAVELIN Renal 101 and IMmotion151 trials, where ccRCC tumors with an LB phenotype (HB.DOWN) exhibited an inflamed tumor microenvironment and responded more favorably to immune checkpoint inhibition.

**Figure 5.**
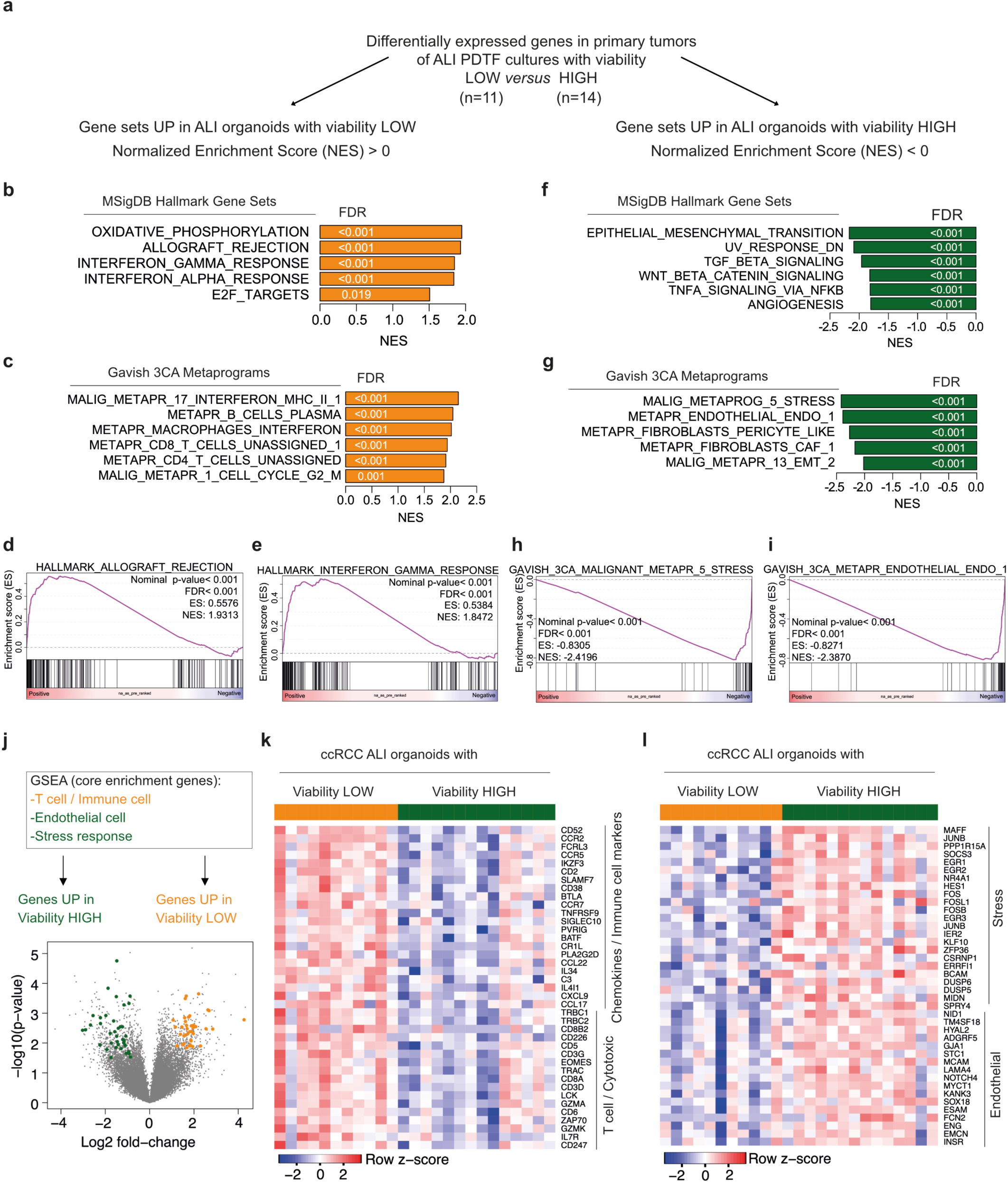
High T cell and chemokine transcript levels associate with reduced viability of ccRCC ALI cultures. **(a)** Outline of transcriptomic comparisons of primary tumors corresponding to ccRCC ALI cultures with low (n=11) versus high (n=14) viability. **(b-c)** Top enriched gene sets in low-viability ccRCC ALI cultures using the (b) Hallmark and (c) Gavish Metaprograms gene set collection from the Molecular Signature Database (MSigDB) for GSEA. NES: normalized enrichment score; FDR: false-discovery rate. **(d-e)** Individual GSEA plots for the indicated gene sets from the Hallmark gene set collection. **(f-g)** Same analyses as described in b-c, but for top enriched gene sets in high-viability ALI cultures. **(h-i)** Individual GSEA plots for the indicated gene sets from the Gavish Metaprograms gene set collection. **(j)** Volcano plot showing differentially expressed genes in primary tumors corresponding to ccRCC ALI cultures with low versus high viability. T cell, immune cell (up in low-viability ALI cultures), and endothelial cell, stress response (up in high-viability ALI cultures) genes are highlighted in orange and green. **(k-l)** Heatmaps showing expression of (k) T cell and chemokine genes, and (l) endothelial cell and stress response genes across all samples.

### Low-branching vessels shape immune cell-rich niches

To further characterize the differences in the TME composition, we conducted high-plex immunofluorescence imaging using the PhenoCycler platform, also known as co-detection by indexing (CODEX)^31,32^. We analyzed full-sections of exemplary primary tumors corresponding to high- and low-viability ALI cultures, the latter with a higher proportion of LB and SN vascular patterns (**Fig. 6a**). Briefly, CODEX employs staining of tissue sections with a panel of antibodies that are conjugated with DNA oligos (barcodes) and subsequent imaging by iterative rounds of hybridization and de-hybridization of complementary fluorophore-tagged DNA oligos (reporter probes)^33^. For image processing, segmentation and cell type annotation, we used the SPACEc workflow^34^. We established a comprehensive antibody panel for the detection and phenotyping of tumor cells, immune cell subtype, endothelial cells and pericytes (**Fig. 6b**, Supplementary Fig. 3 and Supplementary Table 9). In the cases with higher content of LB and SN vascular patterns (samples I and II), we observed dense immune infiltrates along the low-branching vessels, in particular CD8^+^ and CD4^+^ T cells as well as HLA-DR^+^ antigen-presenting myeloid cells (APC) (**Fig. 6c**). Tumors with a predominant HB vascular pattern (Samples III-VI) showed a uniform distribution of immune cells with macrophages being the most frequent immune cell subtype. In conclusion, we identified T cell and APC-rich low-branching vessel-immune cell niches as spatial feature of ccRCC associated with low viability of ALI cultures under IL-2-rich culture conditions, strongly arguing for enhanced intrinsic anti-tumor immunity.

**Figure 6.**
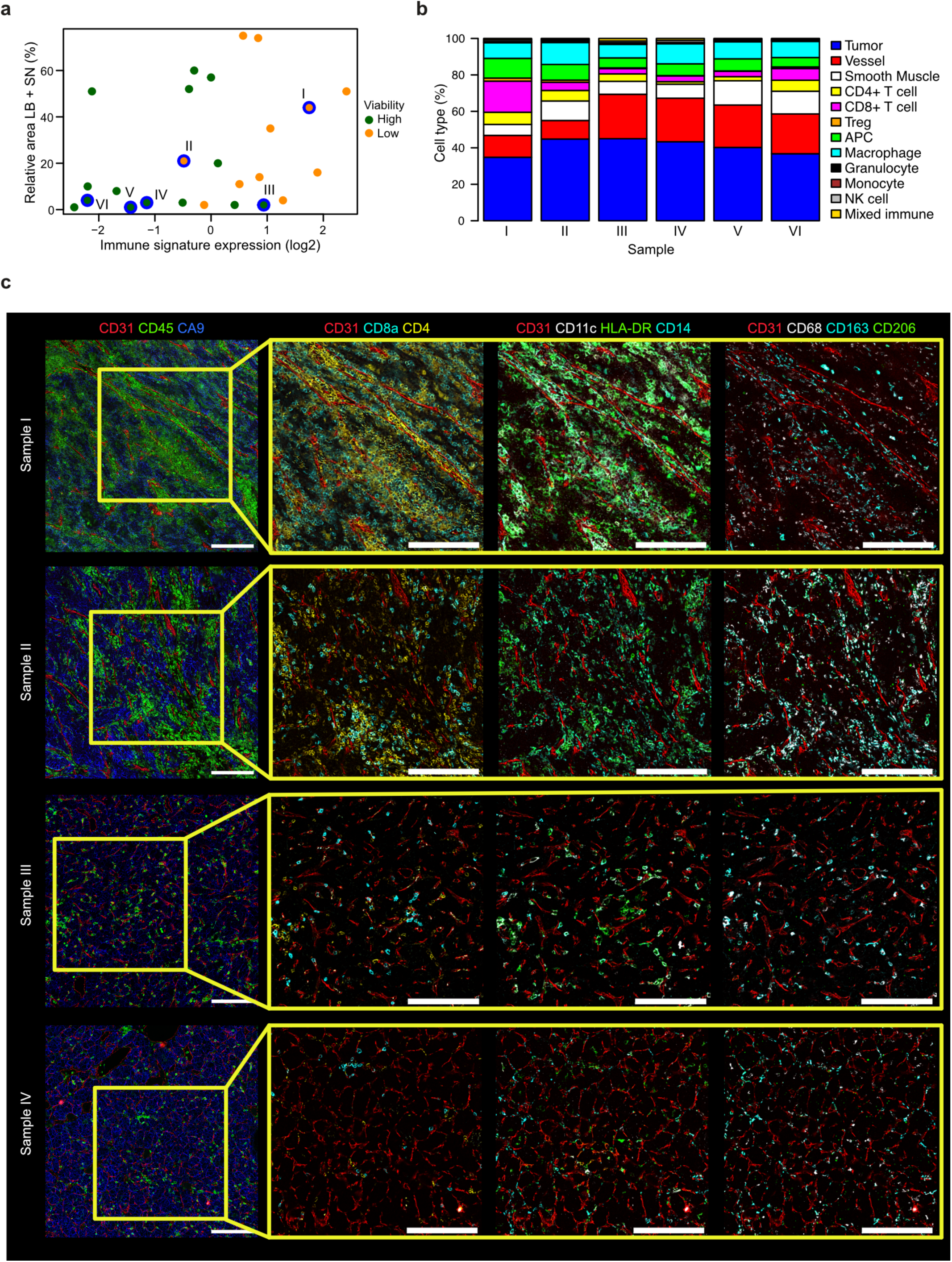
Multiplex immunofluorescence imaging identifies low-branching vessels as T cell and antigen-presenting cell rich niches in the tumor microenvironment of ccRCC. **(a)** Scatter plot integrating the information of immune signature expression (median centered), relative area of low-branching (LB) and sinusoid (SN) vascular pattern, and viability status of ccRCC ALI cohort profiled for mRNA expression (n=25). Cases further analyzed by multiplex immunofluorescence are highlighted by blue circles and numbered by Roman numerals. **(b)** Stacked barplot showing the cell type composition based on the cell type assignment and the antibody panel used for cell phenotyping. **(c)** Exemplary images showing low-branching vessel-immune cell niches in samples I and II, and homogeneously distributed immune cells in tumors with a predominant high-branching vessel architecture (samples III and IV). Left panels: Scale bars represent 200 µm. Right panels: Scale bars represent 200 µm.

## Discussion

We established PropSegNet, a learning-based algorithm for unbiased, automated segmentation of CD31-stained IHC images, classifying vascular patterns into high-branching (HB), low-branching (LB), and sinusoidal (SN) types. Immunohistochemical detection of CD31 and other endothelial markers is widely available in routine pathology laboratories, and when combined with automated image analysis, it enables the development of potential biomarkers based on vessel architecture. Conceptually, integrating morphological information by vascular pattern detection is distinct from microvessel density quantification^9,10^, which primarily counts endothelial cells. Recently, Jasti et al. developed a deep learning model based on H&E stains to predict the RNA-based angioscore that was shown to correlate with responsiveness to anti-angiogenic therapy^35^, but different vascular patterns were not addressed in that study. The work by Jasti et al. demonstrates the feasibility of using H&E stained ccRCC tissue sections to derive vascular networks^35^, clearly a perspective for the further development of PropSeqNet.

Our multiplexed immunofluorescence imaging results supported the idea that vessel architecture provides additional information, as low-branching vessels emerged as regions with dense immune cell infiltration, in particular T cells and MHC class II positive antigen-presenting cells. Thus, we propose that low-branching vessel-immune cell niches (LBVIN) constitute key microenvironmental features in ccRCC associated with enhanced anti-tumor immunity. Notably, several recent studies have underscored the importance of tertiary lymphoid structures (TLS), demonstrating that their presence in ccRCC correlates with responsiveness to ICI therapy^36,37^. When evaluated together, TLS and LBVIN may enhance the prediction of immune checkpoint inhibitor responses and enable refined patient stratification.

Indeed, the translational potential of our findings is also supported by re-analyses of the JAVELIN Renal 101 and IMmotion151 trial datasets^17,19^. In these cohorts, tumors with a predominant LB vascular phenotype (reflected as HB-signature downregulation) were associated with an inflamed microenvironment and demonstrated improved responses to combinations of ICI and VEGF signaling inhibitors. In contrast, tumors displaying a predominant HB vascular phenotype were characterized by a non-inflamed microenvironment and patients did not experience a PFS benefit from the combination therapy. On the other hand, these patients demonstrated an overall more favorable prognosis arguing that the HB vascular phenotype represents a morphological correlate of less aggressive tumors. This concept is further substantiated by the robust co-expression of proximal tubular cell lineage genes, consistent with a higher degree of tumor cell differentiation at the molecular level. Thus, vascular patterns may serve as both predictive and prognostic biomarkers in ccRCC, thereby complementing existing grading schemes^1,3^.

Mechanistically, the enrichment of T cell and chemokine-related transcripts in low-viability ALI cultures implies that tumor-intrinsic immunity, amplified by IL-2 in the standard ALI protocol described by Neal et al.^29^, drives increased cytotoxicity and tissue attrition *in vitro*. This is supported by a recent study, which demonstrated that a novel fusion protein targeting IL-2 to CD8^+^ T cells effectively reinvigorated tumor-intrinsic immunity in patient-derived tumor fragments (PDTF) cultured in vitro^38^. Given that primary tumors of low-viability ALI cultures were enriched for LB and SN vascular patterns, the findings are also consistent with our transcriptomic analysis of the JAVELIN Renal 101 and IMmotion151 cohorts, which showed that ccRCC with an inflamed LB vascular phenotype responded more favorably to ICI.

The air-liquid-interface (ALI) method was previously described as an improved approach for patient-derived organoid modelling, with the advantage of partially preserving the tumor microenvironment including stroma, immune cells and extracellular matrix^29,39^. Improved oxygenation relative to conventional submerged cultures may also reduce tissue necrosis due to hypoxia^29,40,41^. The ALI protocol involves culturing small pieces of patient-derived tumor fragments (PDTF), in contrast to classical organoids derived from single cell suspensions of cancer cells. PDTF platforms were successfully used to study T cell activation by immune checkpoint inhibitors in vitro and dissect the variability of responses^42,43^. Thus, recent advances in different PDTF culture techniques hold great promise to test novel immunotherapeutics and facilitate their translation into clinical trials^38^.

Nevertheless, we experienced several challenges and limitations in our work with ALI cultures of ccRCC PDTF. Besides tissue availability and fragment size, tumor cell content and debris introduced a high degree of variability in our study, which confounded our initial attempts to assess tumor tissue viability by ATP- or LDH-based assays, and flow cytometry (live-dead stain). For example, flow cytometry was biased for immune cell detection, whereas larger ccRCC cells appeared fragile and underrepresented. Based on these experiences, we adopted a conservative approach by fixation, serially sectioning and evaluation of the viable tissue area in each section. On the other hand, this strategy limited tissue availability for complementary assays. Furthermore, our previous pilot study revealed that cultured tissue fragments must be relatively small^30^, in line with other studies working with PDTF cultures^29,38,42^, which limited the robust assessment of vascular patterns within these samples, and consequently we focused our analysis on the matched primary tumor tissue sections.

In summary, our study provides a comprehensive evaluation of the vascular architectures in ccRCC by integrating automated image analysis, transcriptome profiling, multiplexed immunofluorescence, and PDTF modeling. Our findings demonstrate that the vessel architecture in ccRCC not only reflects the lineage characteristics of tumor cells but also co-evolves with features of the immune microenvironment, an observation that may have important implications for therapy stratification in combination with emerging blood- and imaging based markers^44–47^. Moreover, the automated quantification of vascular patterns using PropSeqNet is a potentially widely applicable method, but its predictive value for patient stratification will require validation in prospective studies, which may be facilitated by adapting the approach to H&E-based tissue sections^35^.

## Methods

### Immunohistochemistry (IHC)

Staining was performed according to the protocols of the routine laboratory at the Institute of Pathology, University Hospital Bonn (Germany). The staining for CD31 (mouse anti-human CD31, clone JC/70A, dilution 1:100, Dako) was performed on the Medac autostainer 480S (Medac), using a C-DPVB 500 HRP–detection system.

### Learning-based algorithm PropSegNet

A detailed description of the development of PropSegNet (Proportion Segmentation Network) is provided in Supplementary Methods. Patient characteristics of the cohorts used for training and validation of PropSeqNet is provided in Supplementary Table 1. The Ethics Committee at the University Hospital Bonn, University of Bonn, Bonn, Germany gave ethical approval to this work including AI-based methods to detect tumor growth pattern (Az. 2024-312-BO).

### RNA and DNA isolation from FFPE tissue

FFPE tissue blocks were cut using a microtome to and placed onto multiple slides to obtain enough tissue for RNA and DNA isolation. The slides were placed on a prewarmed heating block to soften the paraffin and facilitate the extraction of material from the slides. The tumor areas were scraped from the slides using a scalpel and transferred to PCR-Safe 2 mL reaction tubes (Eppendorf) and stored at 4°C until further processing. For isolation of RNA and DNA from the formalin fixed and paraffin embedded tissue, the Qiagen AllPrep DNA/RNA FFPE Kit (50) was used according to manufacturer protocol. Concentration of RNA in the samples was determined using a NanoDrop and samples were submitted to the NGS Core Facility for 3’mRNA sequencing. The libraries are prepared by using the QuantSeq FWD 3’-mRNA-Seq Kit from Lexogen GmbH (Austria). The sequencing is done on the NovaSeq 6000 with a read length of 1 x 100bp and in most cases an average of 10 M raw reads per sample.

### Patient samples (ccRCC ALI cohort, FFPE cohorts)

The ALI cohort included 30 samples from clear cell renal cell carcinoma from patients who underwent complete or partial nephrectomy between 2019 and 2020 in the Department of Urology at the University Hospital in Bonn (Germany). All patients gave informed consent for biobanking. The Ethics Committee at the University Hospital Bonn, University of Bonn, Bonn Germany gave ethical approval for the work involving organoids as well as biomarker analyses using fresh-frozen or FFPE archived tissues (immunohistochemistry, transcriptome analyses, mutations) (EK417/17; EK 233/20; EK 285/21). The clinicopathological characteristics of the patients are summarized in Supplementary Table 6.

### Cultivation of the air-liquid-interface patient-derived tumor fragments

Six inserts (0.4 μm diameters, Milicell-CM, Milipore) per case were coated with a mixture of collagen I, Ham’s F12 and reconstitution buffer (mixing ratio 8:1:1) per well. The inserts were placed into a 6-well plate and incubated for 30 minutes at 37°C to solidify the collagen. The tumor samples were transferred into DPBS (1 x Dublecco’s Phosphate Buffered Saline, Gibco) on ice and manually minced into small pieces (∼0.5-1 mm^3^), afterwards dissociated in the GentleMACS Octo tissue dissociator (Milteniy Biotec). The material was washed with ADMEM-F12 (Thermo Fisher) containing normocin and amphotericin B and mixed with collagen I, Ham’s F12, reconstitution Buffer mixture. 1ml of the tumor-collagen mixture was transferred to the prior prepared wells. 1 ml of growth factor media containing ADMEM-F12, supplemented with 50% Wnt3a, R-spondin 1 conditioned medium from L-WRN feeder cells (obtained from ATCC; CRL-3276), with 1 mM HEPES (Thermo Fisher), 1 x Glutamax (Thermo Fisher), 10 mM nicotinamide (Sigma), 1 mM N-acetylcysteine (Thermo Fisher), 1x B27 without vitamin A (Thermo Fisher), 0.5 mM A83-01 (Sigma), 1x penicillin/streptomycin (Thermo Fisher), 10 nM gastrin (Sigma), 10 mM SB-202190 (Peprotech), 50 ng ml^-1^ EGF (Sigma), 25 ng ml^-1^ noggin (Invitrogen), 100 mg ml^-1^ normocin and IL-2 (600 UI ml^-1^, Peprotech) were added into the outer part of the wells and incubated at 37°C. The media was substituted twice a week and tumor growth observed regularly under the microscope. ALI PDOs were split 1:2 once per two to three weeks, depending on the growth rate. For splitting, we removed the media, added a mixture of PBS and 200 Units of collagenase IV (Rockland) into the insert and incubated at 37°C until the collagen was fully dissociated. The tissue fragments were washed, the collagenase activity was inhibited with trypsin-EDTA (0.25%). Subsequently, the tissue fragments were then re-plated as described above into new inserts.

### Fixation and tissue processing of ccRCC ALI cultures

The ccRCC ALI cultures within the collagen gels were cut out with a scalpel, fixed in buffered formaldehyde, transferred into HistoGel (Richard-Allan Scientific) and embedded in paraffin. Since the tissue fragments are very small, FFPE embedded material was cut in 10-30 step sections with 2 µm each and stained with Haematoxylin and Eosin (H&E) to visualize the tumor tissue and to determine the degree of necrosis by board-certified pathologist with more than 20 years of work experience specialized on renal pathologies. For quantifying the treatment response, as necrotic areas per total area of organoids, we scanned the slides using a Leica Aperio GT 450DX scanner and analyzed the pictures with the open source QuPath-0.4.3.

### RNA isolation and sequencing from fresh-frozen tissue (ccRCC ALI cohort)

Fresh-frozen tissue, previously stored at -80°C, was added to a Miltenyi gentleMACS M Tube containing 600 µL of buffer RLT (Qiagen RNeasy Mini Kit) at room temperature. The samples were then subjected to the RNA_02 program on the gentleMACS Octo Dissociator followed by centrifugation at 2000 x g for 1 min. RNA isolation was then performed using the RNeasy Mini Kit, following the manufacturer protocol. Concentration of RNA in the samples was determined using a NanoDrop and samples were submitted to the NGS Core Facility for 3’mRNA sequencing using the forward QuantSeq 3′mRNA-seq Library Prep Kit for Illumina (Lexogen GmbH, Austria) according to the manufacturer’s protocol.

### Differential gene expression and gene set enrichment analysis (GSEA)

Differential gene expression analysis (ccRCC ALI cohorts) was performed using the limma functions ‘lmFit’, ‘eBayes’ (eBayes moderated t-test statistics) and ‘topTable’. The contrast design was high-viability versus low-viability. Gene set enrichment analysis (GSEA) was performed using the Java-based stand-alone version downloaded from https://www.gsea-msigdb.org/gsea/index.jsp. The Hallmark and Gavish Metaprograms gene set collections were obtained from the Molecular Signature Database (MSigDB) provided through the aforementioned link or accessed directly via the Java-based stand-alone version of GSEA. Single cell type signatures of the Human Protein Atlas (HPA) project were download from https://www.proteinatlas.org/ and compiled into a gmt file compatible with the GSEA software (Supplementary Table 5). The pre-ranked gene list mode was used for the analyses with 1000 permutations and default settings. Vector graphics compatible GSEA plots were re-generated with a modified version of the R-based function ‘replotGSEA’ obtained from the following github repository: https://github.com/PeeperLab/Rtoolbox/blob/master/R/ReplotGSEA.R.

### Correlation of vascular patterns with gene expression

Relative area (rel_area) of high-branching (HB), low-branching (LB) and sinusoid (SN) vascular patterns classified by PropSegNet were correlated with each gene of the gene expression matrix (expr_matrix) using the R function ‘apply’: apply(expr_matrix, 1, cor, rel_area, method = “Spearman”). Cutoffs for rho were arbitrarily chosen (HB: rho > 0.4; LB: rho > 0.3, SN: rho > 0.3) to obtain gene signatures of recommended size for downstream analyses like GSEA.

### Protein expression in normal kidney (HPA)

Images were downloaded from the Human Protein Atlas (HPA) website. Screenshots with links, antibody and HPA patient sample identifiers are provided in Supplementary Figure 2.

### Access to RNAseq data from the JAVELIN Renal 101 cohort

Normalized RNA-seq data and clinical sample annotation were downloaded from the supplementary data section provided with the original publication by Motzer et. al.^20^. Data were imported into the R/Rstudio computing environment for re-analyses.

### Access to RNAseq data from the IMotion151 cohort

Access to the following IMotion151 datasets has been granted by the Data Access Committee (DAC): EGAD00001006616 (FMI data: somatic alteration calls summarized at the gene level for 715 patients profiled by FoundationOne), EGAD00001006617 (Demographic, histology, PDL1 IHC, TMB and outcome data, PFS and ORR for 836 patients), EGAD00001006618 (Processed RNAseq data: log2(TPM + 1) transformed counts for the 823 tumor samples profiled by RNAseq) and EGAD00001006619 (Raw FASTq files for tumors from the 823 patients profiled by RNAseq). Processed RNAseq data were used for the transcriptomic analyses. Data were imported into the R/Rstudio computing environment for re-analyses.

### Survival analyses (JAVELIN 101 and IMotion151 cohorts)

Expression of the high-branching (HB) gene signature per sample was determined by averaging (mean) log2-transformed gene expression values. We then followed the same analytical strategy as described by Motzer et al.^20^, dividing the entire cohort by an unbiased median classifier into “high-branching signature UP” and “high-branching signature DOWN”. The latter subgroup is presumed to be enriched with tumors with a sinusoid and low-branching vascular pattern, in particular. Subsequently, we separated the subgroups by the different treatment arms of the JAVELIN Renal 101 and IMotion151 cohorts, respectively. Survival analyses were performed using standard approaches available through respective R packages. Briefly, median progression-free survival (PFS) was determined with the function surv_median from the R package survminer. Logrank-test was performed with the function logrank_test from the R package coin. Hazard ratios (HR) and confidence intervals were determined with the function coxph from the R package survival.

### Comparison to NMF cluster

Assignment of samples from the IMotion151 cohort to NMF clusters described by Motzer et al.^17^ was retrieved from the EGAD00001006617 dataset. Association with high-branching signature UP or high-branching signature UP subgroups were determined by two-sided Fisher’s exact test using the function fisher.test from the basic R package stats.

### Amplicon sequencing (ccRCC driver mutations)

The DNA concentration was determined on a Quantus™ fluorometer using the QuantiFluor® ONE ds DNA System (Promega). Generation of the sequencing library was performed using a QIAseq^TM^ targeted DNA custom panel (Qiagen) with an input of 40 ng DNA according to the manufacturers protocol. Amplification products were subjected to next generation sequencing on an Illumina MiSeq sequencer (Illumina). Sequencing data were analyzed for genomic variants using the CLC Genomics Workbench/Server (Qiagen Bioinformatics), usind the following filtering criteria: Alt.Variant Freq ≥ 1%; for/rev balance ≥ 0.1; QUAL ≥ 200; AvQuality ≥ 38; Read pos. test probability ≥ 1*10^-6^; Read dir. test probability ≥ 1*10^-6^. Variants were then manually filtered for artifacts, synonymous variants and assumed germline variants and relevant somatic variants with an alternative allele frequency ≥ 5% were identified. For classification and interpretation of somatic variants the following databases were used: *dbSNP, COSMIC, OncoKB, ClinVar, cBioPortal* in their respective current versions. Targeted regions of QIAseq targeted DNA custom gene panel are provided in **Supplementary Table 8**.

### Buffers and solutions for multiplexed immunofluorescence

TCEP-reducing solution: 2.5 mM TCEP (Sigma, 646547) and 2.5 mM EDTA pH 8.0 (Invitrogen, AM9261) in ddH_2_O, pH 7.0. Buffer C: 150 mM NaCl (Carl Roth, 9265.2), 2 mM Tris stock solution (Carl Roth, AE15.3), pH 7.2, 1 mM EDTA, and 0.02% w/v NaN_3_ (AppliChem, A14300,1000) in ddH_2_O. High-salt PBS: 900 mM NaCl in 1× DPBS (Gibco, 14190-094). CODEX® antibody stabilizer solution: 0.5 M NaCl, 5 mM EDTA, and 0.02% w/v NaN_3_ in PBS antibody stabilizer solution (CANDOR Biosciences GmbH, 131125). Staining solution 1 (S1): 5 mM EDTA, 0.5% w/v bovine serum albumin (BSA, Carl Roth, 8076.3) and 0.02% w/v NaN_3_ in 1× DPBS, stored at 4 °C. Staining solution 2 (S2): 61 mM NaH_2_PO_4_ (Sigma, S0876), 39 mM NaH_2_PO_4_ · H_2_O (Sigma, S9638), 250 mM NaCl in a 1:0.7 v/v solution of S1 and double-distilled H_2_O (ddH_2_O); final pH 6.8–7.0, stored at 4 °C. Staining solution 4 (S4): 0.5 M NaCl in S1, stored at 4 °C. Blocking buffer: S2 buffer containing B1 (1:20), B2 (1:20), B3 (1:20), and BC4 (1:15), stored at 4 °C. Blocking reagent 1 (B1): 1 mg/ml mouse IgG (Sigma, I5381) in S2, stored at 4 °C. Blocking reagent 2 (B2): 1 mg/ml rat IgG (Sigma, I4121) in S2, stored at 4 °C. Blocking reagent 3 (B3): sheared salmon sperm DNA (Invitrogen, AM9680), 10 mg/ml in H_2_O, stored at 4 °C. Blocking component 4 (BC4): Mixture of 57 non-modified oligonucleotides (Biomers) at a final concentration of 0.05 mM each in TE buffer (Sigma, 93302), stored at 4 °C (Supplementary Data <u>5</u>). BS3 fixative solution: 200 mg/ml BS3 (ThermoFisher, 21580) in DMSO from a freshly opened ampoule (Sigma, D2650-5x5ML), stored at 20 °C in 3 µl aliquots. H2 buffer: 150 mM NaCl, 10 mM Tris pH 7.5, 10 mM MgCl_2_ · 6 H_2_O (Carl Roth, 2189.1), 0.1% w/v TritonTM X-100 (Sigma, X-100) and 0.02% w/v NaN_3_ in ddH_2_O. Plate buffer: H2 buffer containing DAPI nuclear stain (1:300, Biolegend, 422801) and 0.5 mg/ml sheared salmon sperm DNA. Fluorescent oligonucleotide stock solution (Biomers): 100 µM Fluorescent oligonucleotide dissolved in 1× TE buffer, stored in the dark at −20 °C. Fluorescent oligonucleotide working solution: Fluorescent oligonucleotide stock solution diluted 1:10 in 1× TE buffer, stored in the dark at 4 °C. Plate Buffer: H2 buffer containing DAPI nuclear stain (1:300) and 0.5 mg/ml sheared salmon sperm DNA.

### Multiplexed immunofluorescence of tumor tissue

All tumor samples were obtained following informed consent as part of SOC surgical procedures. All patients had consented to in-depth analyses of tissue. FFPE tumor samples were sliced by standard procedures at 3 µm slice thickness and adhered onto poly-L-lysine-coated coverslips. Antibody conjugation, tissue staining, and mIF imaging were performed (with modifications) as described elsewhere^33^. In short, purified, carrier-free antibodies were conjugated to maleimide-modified oligonucleotides (Biomers), upon reduction in presence of TCEP-reducing solution. Maleimide-modified oligonucleotides were first dissolved in 1× DPBS, then added to the reduced antibody and incubated at room temperature for two hours in a 2:1 (w/w) ratio with the antibodies. Next, the conjugated antibodies were washed in high-salt PBS three times and then eluted by centrifugation at 3000 × *g* for 2 min in the CODEX® antibody stabilizer solution. The conjugated antibodies were stored at 4 °C until usage. Prepared FFPE tissues were baked at 55 °C for 30 min, and rehydrated by immersion in fresh xylene, twice, for 5 min and in descending concentrations of ethanol, each step for 5 min (100% twice, 95% twice, 70%, ddH2O twice). Heat-induced epitope retrieval was performed using 1× Dako target retrieval solution, pH 9 (Agilent, S2367) at high pressure, for 20 min. Tissues were then washed for 10 min in 1× TBS IHC wash buffer with Tween 20 (ThermoFisher, 28360). Tissues were blocked for 1 h at room temperature using 100 µl of blocking buffer. Conjugated antibodies were added to the blocking buffer, concentrated through a 50 kDa Amicon Ultra Filter, and resolved in the blocking buffer. Tissues were incubated with the antibody staining solution in a humidity chamber overnight at 4 °C. Details on the antibodies and DNA oligo barcodes are provided in Supplementary Table 9. After staining, tissues were washed twice in S2 buffer and fixed with a three-step fixation process. First, tissues were fixed in S4 containing 1.6% paraformaldehyde (Electron Microscopy Science, 15710-S) for 10 min, followed by a 15 min-long incubation in 100% ice-cold methanol (Sigma, 34860-1L-R) for 5 min, and a final fixation with BS3 fixative solution dissolved in 1× PBS at room temperature for 20 min. Tissues were stored in S4 in a six-well plate at 4 °C for up to 2 weeks, or further processed for imaging. 400 nM fluorescent oligonucleotide working solution was aliquoted in Corning^TM^ black 96-well plates (Merk, CLS3925-100EA) in 250 μl of plate buffer, according to the multi-cycle reaction panel. Image acquisition was performed on Zeiss Axio Observer 7 microscope equipped with a Colibri 7 LED Light source (Carl Zeiss), and a Prime BSI PCIe camera (Teledyne Photometrics). Imaging cycles were performed using an Akoya Phenocycler™ instrument and CODEX® instrument manager software (Akoya Biosciences). Automated images were acquired with the Plan-Apochromat 20×/0.8 M27 (*a* = 0.55 mm) objective (Carl Zeiss), and the imaging pipeline was controlled by a focus strategy with autofocus for each support point created, with a three z-stack image with a distance of 1.5 μm. DAPI (1:300 final concentration) was imaged in each cycle at an exposure time of 20 milliseconds and LED intensity of 40%.

### Multiplex immunofluorescence image processing and analyses

Image selection criteria: Before analyzing the images staining, imaging and tissue quality were assessed by manual inspection of the images. Images that were out of focus or contained only low-quality destroyed samples were excluded from the analysis. Markers with insufficient staining quality were excluded individually.

To identify the cell types in the RCC dataset spatial analysis was executed using the SPAECc workflow^34^ available at https://github.com/yuqiyuqitan/SPACEc. Before preprocessing the images with SPACEc, they were segmented using Instanseg^48^ (available in GitHub:: https://github.com/instanseg/instanseg?tab=readme-ov-file) via QuPath v0.6.0-rc3^49^ (available in github: https://github.com/qupath/qupath/releases/tag/v0.6.0-rc3).

Segmentation was performed as a tiled approach (1024 x 1024 px tile size) over tumor regions, excluding the tumor capsule or surrounding tissue (manual region of interest selection). Cell size and mean intensities for all fluorescent markers were measured per cell mask via QuPath. Segmentation quality was assessed by visual inspection. After segmentation, cells that were too small or had too low DAPI intensity were removed from the analysis to exclude segmentation artifacts. For that, we started with the lowest 1% and adjusted the threshold accordingly based on plotting the size and DAPI intensity distribution. Subsequently, the intensities for every marker were normalized per image via Z-score normalization^50^. After that, we removed cells that were positive for too many markers by thresholding the top 1% of positive Z-score count and Z-score sum^34^. Cells were then clustered into four major groups by GPU-accelerated Leiden clustering (https://zenodo.org/records/12533399) on phenotypic markers (’FOXP3’, ’GNZB’, ’CD4’, ’CD31’, ’CD8a’, ’CD3e’, ’CD68’, ’CA9’, ’CD45’, ’HLADR’, ’CD14’, ’CD56’, ’CD15’, ’CD11c’, ’CD20’, ’CD11b’, ’CD163’, ’Perforin’, ’CD206’, ’aSMA’) and manual inspection of dominating markers in the clusters. Major groups were differentiated into tumor (CA9+), immune (CD45+) stroma (CD31+, aSMA+), and noise (no marker expression). These groups were then re-clustered using Leiden clustering. Cell types were assigned based on the dominating phenotypic markers in each cluster and manual annotation. Subsetting and re-clustering were repeated three to four times (multiple major rounds + re-clustering of single groups if necessary) to derive the shown cell type labels (supplemental figure 3). During the clustering steps cells were excluded that were identified as noise. We classified cells as noise if they were negative for all markers or positive for unspecific markers due to high staining background and not captured during the prior filtering steps. Before removing cells their identity as noise was confirmed by plotting the centroids onto the original image using TissUUmaps^51^. Additionally, we manually excluded areas where the tissue or staining quality was insufficient or which were outside the tumor (remaining parts of the capsule). After successful annotation, we calculated the respective cell type percentage per image. The derived dataset includes six regions and 2924723 single cells.

## Supporting information

Supplementary_Methods_prop_segNet

Supplementary_Table_1_PropSegCohort

Supplementary_table_4_VP_gene_signatures

Supplementary_Table_5_HPA_Single_Cell_Types_Ensembl

Supplementary_Table_6_ALI_cohort_pat_characteristics

Supplementary_Table_8_QiagSeq

Supplementary_Table_2_3_advanced_RCC_characteristics

Supplementary_Table_7_ALI_cohort_summarized_data_rev

## Data Availability

Data availability:
Raw 3'mRNA sequencing data will be available in Zenodo. Processed data from the JAVELIN Renal 101 study can be accessed from the publication by Motzer et al.20. Redistribution of the data from IMotion151 study is not possible17, but data access can be requested from the Genentech data access committee (DAC, EGAC00001001813) via the European Genome-Phenome Archive (EGA): https://www.ega-archive.org/dacs/EGAC00001001813. Other data produced in the present work are contained in the manuscript as supplementary tables.
Code availability:
Code for PropSegNet will be published on acceptance. The SPACEc code for the analyses of multiplex immunofluorescence imaging data is available on GitHub: https://github.com/yuqiyuqitan/SPACEc.34 Standard code for the analyses of multiplex immunofluorescence imaging data and survival analyses will be also available in GitHub.

## Data availability

Raw 3’mRNA sequencing data will be available in Zenodo. Processed data from the JAVELIN Renal 101 study can be accessed from the publication by Motzer et al.^20^. Redistribution of the data from IMotion151 study is not possible^17^, but data access can be requested from the Genentech data access committee (DAC, EGAC00001001813) via the European Genome-Phenome Archive (EGA): https://www.ega-archive.org/dacs/EGAC00001001813.

## Code availability

Code for PropSegNet will be published on acceptance. The SPACEc code for the analyses of multiplex immunofluorescence imaging data is available on GitHub: https://github.com/yuqiyuqitan/SPACEc.^34^ Standard code for the analyses of multiplex immunofluorescence imaging data and survival analyses will be also available in GitHub.

## Acknowledgements

The authors would like to thank Kerstin Fuchs, Carsten Golletz, Susanne Steiner and the laboratory and medical staff of the Institute of Pathology for their great support. We thank Prof. Dr. Hubert Schorle for designing the gene panel for mutations analysis. We would like to thank NIAID NIH BIOART (https://bioart.niaid.nih.gov) and bioicons (https://bioicons.com) for providing public domain icons for figures. We would like to thank the NGS and Bioinformatics Core Facility of the Medical Faculty at the University of Bonn for providing support and instrumentation funded by the Deutsche Forschungsgemeinschaft (DFG, German Research Foundation). M.H. is a member of the CANTAR project, which receives funding from the Netzwerke 2021 program, an initiative of the Ministry of Culture and Science of the State of North Rhine-Westphalia. The sole responsibility for the content of this publication lies with the authors. A.E. and T.P. were funded by the Deutsche Forschungsgemeinschaft under Germany’s Excellence Strategy—EXC-2047/1-390685813 and EXC2151-390873048. M.H. was supported Deutsche Forschungsgemeinschaft under Germany’s Excellence Strategy EXC2151-project ID 390873048. M.I.T and M.H. were supported by a joint grant by the Deutsche Forschungsgemeinschaft - project ID 497667643. M.I.T., A.E., M.H. are members of ImmunoSensation – the immune sensory system (EXC2151).

## Author contributions

Acquisition of data: M.I.T., M.K., L.E., K.B., S.L., R.T., M.C.R.Y., N.P., S.K. Data analysis and visualization: M.I.T., T.K., M.K., N.P., L.E., K.B., R.T. Computation and coding: Y.L., T.P., G. A-E., A.E., T.K., M.H. Learning-based model: Y.L, T.P., G. A-E., E. B-C., A.E. Resources: M.I.T., G.K., M.R., J.E., M.H. Provision of study materials and curated clinical outcome data: V.G., G.K., M.R., M.I.T., J.E., N.K. Review and editing of original draft: M.I.T, V.G., M.E., L.F., F.H., J.K., J.S., G.K., M.R., J.E. Conceptualization and supervision: M.I.T., N.K., A.E., M.H. Writing: M.I.T, N.K., A.E., M.H. All authors read and approved the final manuscript.

## Legends to Supplementary Figures

**Supplementary Figure 1.**
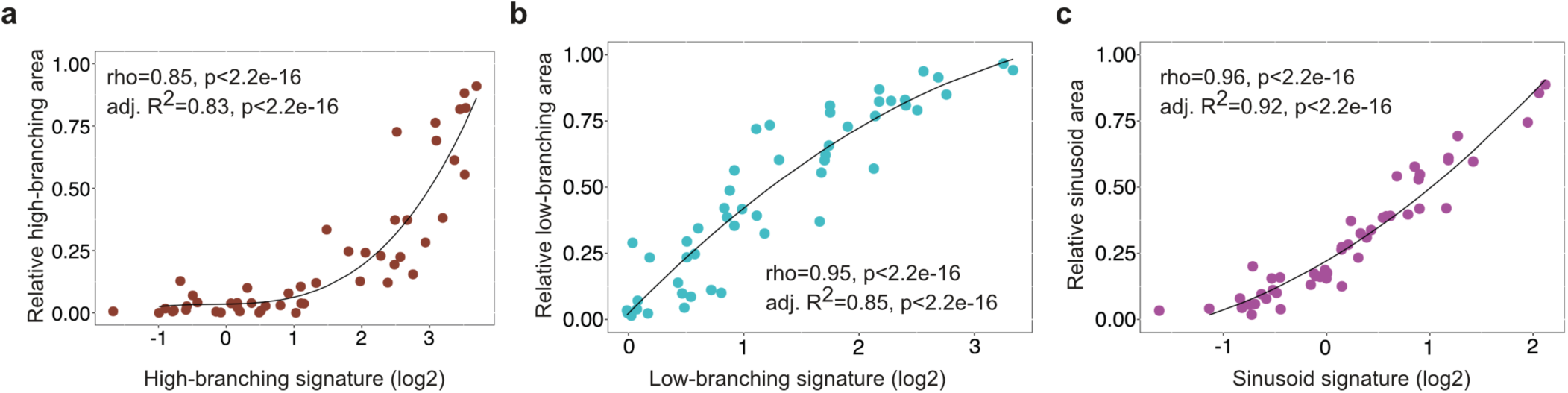
Correlation between gene signature expression and PropSegNet values. Rank correlation of averaged (a) high-branching, **(b)** low-branching, and **(c)** sinusoid transcriptional signature with corresponding relative areas determined by PropSegNet.

**Supplementary Figure 2.**
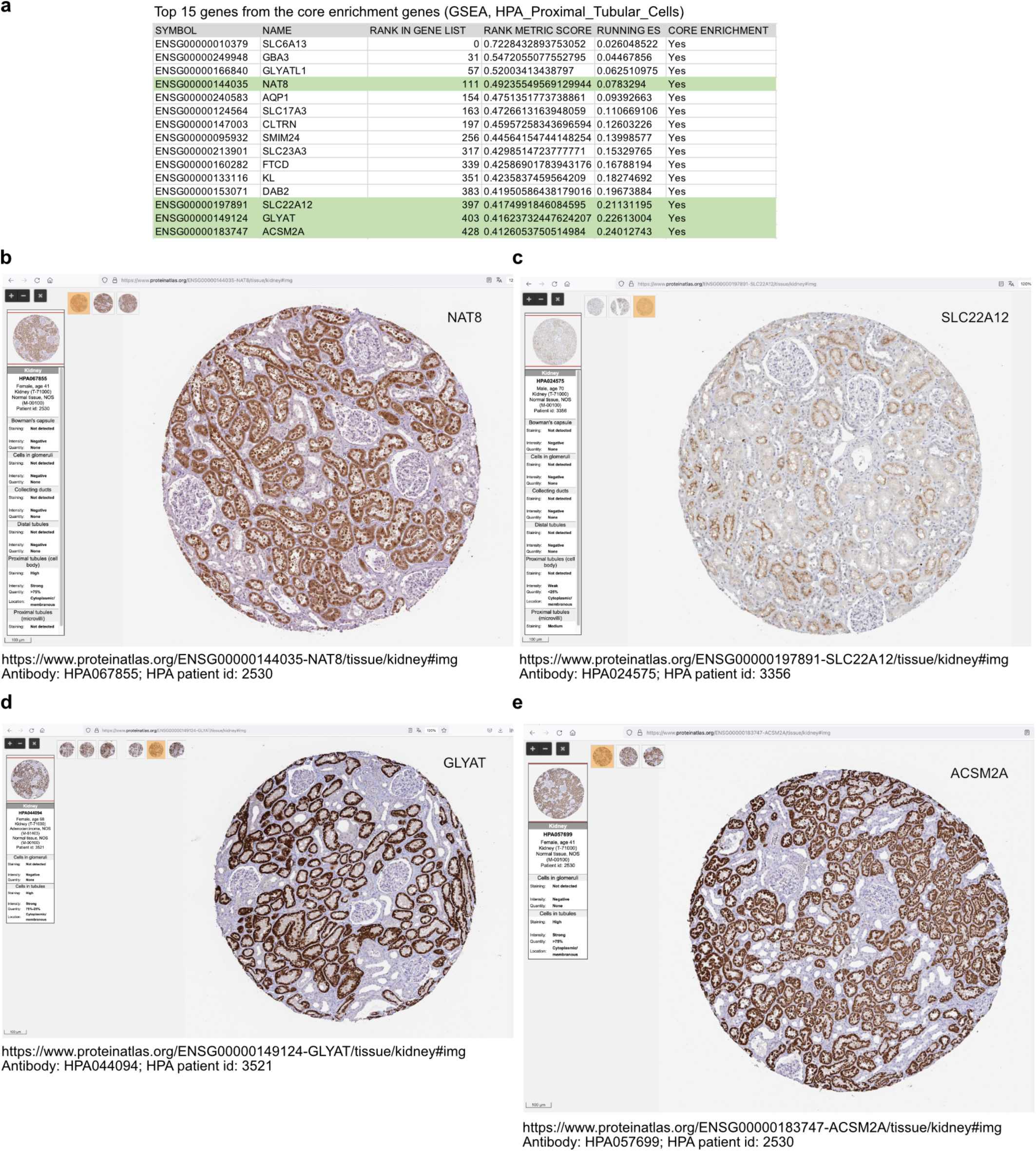
Protein expression encoded by core enrichment genes from the HPA proximal tubular cell gene set. **(a)** Top 15 genes from the core enrichment. Source images, HPA patient id, antibody information and web links from the HPA website for the selected candidate proteins shown in main figure 2: **(b)** NAT8, **(c)** SLC22A12, **(d)** GLYAT, and **(e)** ACSM2A.

**Supplementary Figure 3.**
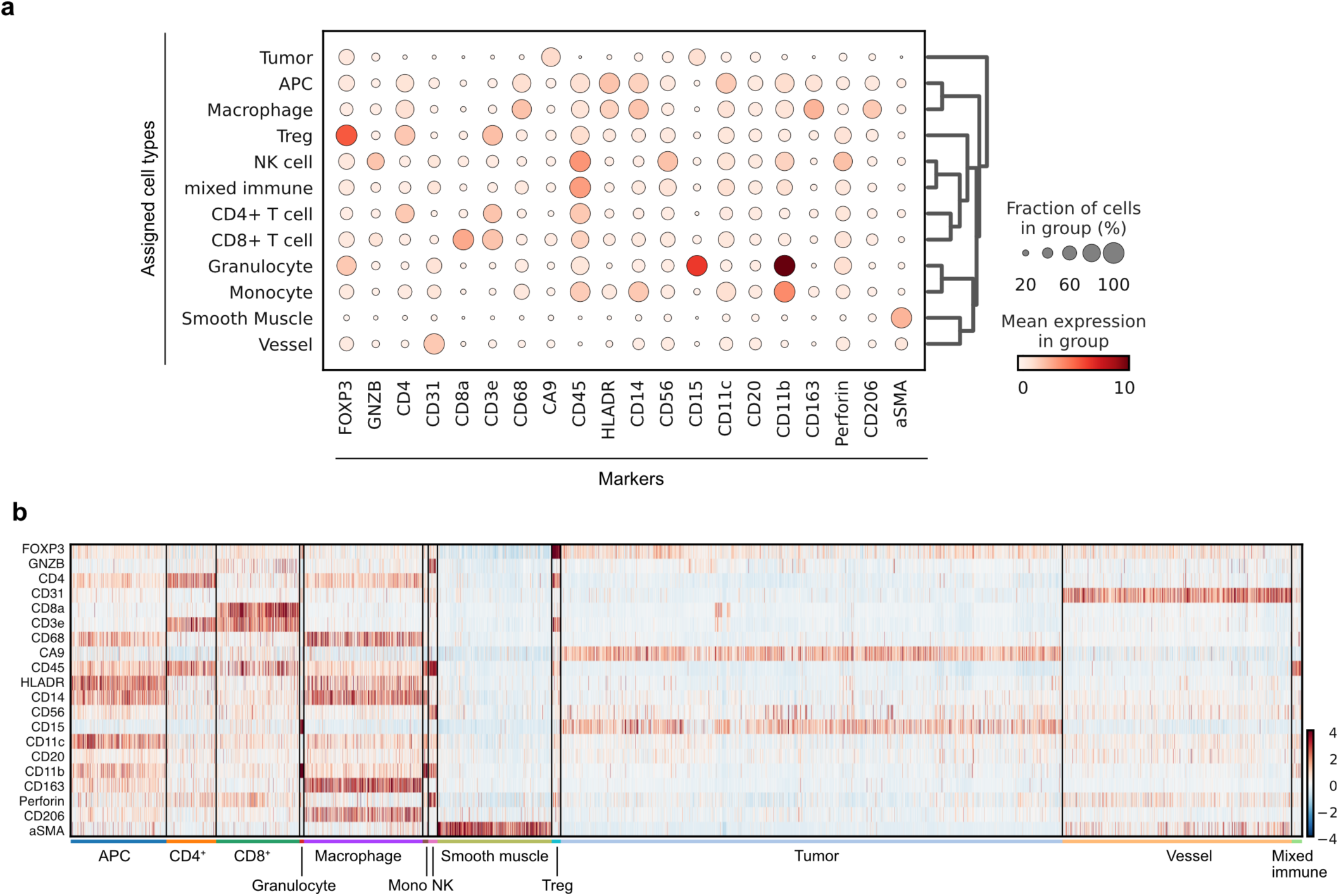
Cell type assignment based on multiplex immunofluorescence imaging data. **(a)** Cell type assignment based on the antibody panel used for multiplex immunofluorescence imaging and mean expression of indicated markers is shown by color coding. **(b)** Heatmap showing expression of markers in cells across data set (z-score transformed).

