## Supplementary_Methods_prop_segNet for "Low-branching vessel architecture shapes immune cell niches and predicts immune responses in renal cancer"

### Supplementary Material

#### Overview of PropSegNet

For the automated segmentation and subsequent classification of patterns in CD31-stained slide images, we propose a novel learning-based algorithm, termed *PropSegNet*. As illustrated in Figure 1, immunohistochemical staining for CD31 in clear cell renal cell carcinoma serves as input to a Swin Transformer-based neural network—a hierarchical transformer architecture utilizing a shifted window mechanism. Notably, only the global proportions of the patterns are provided as ground truth labels during training. As output, the network predicts patch-level proportions corresponding to the structural patterns, and applies test-time augmentation (TTA) to generate pixel-wise segmentation masks. The proportion of each pattern is then computed from the relative area of the corresponding segmentation masks (implementation details are provided in the supplementary material).

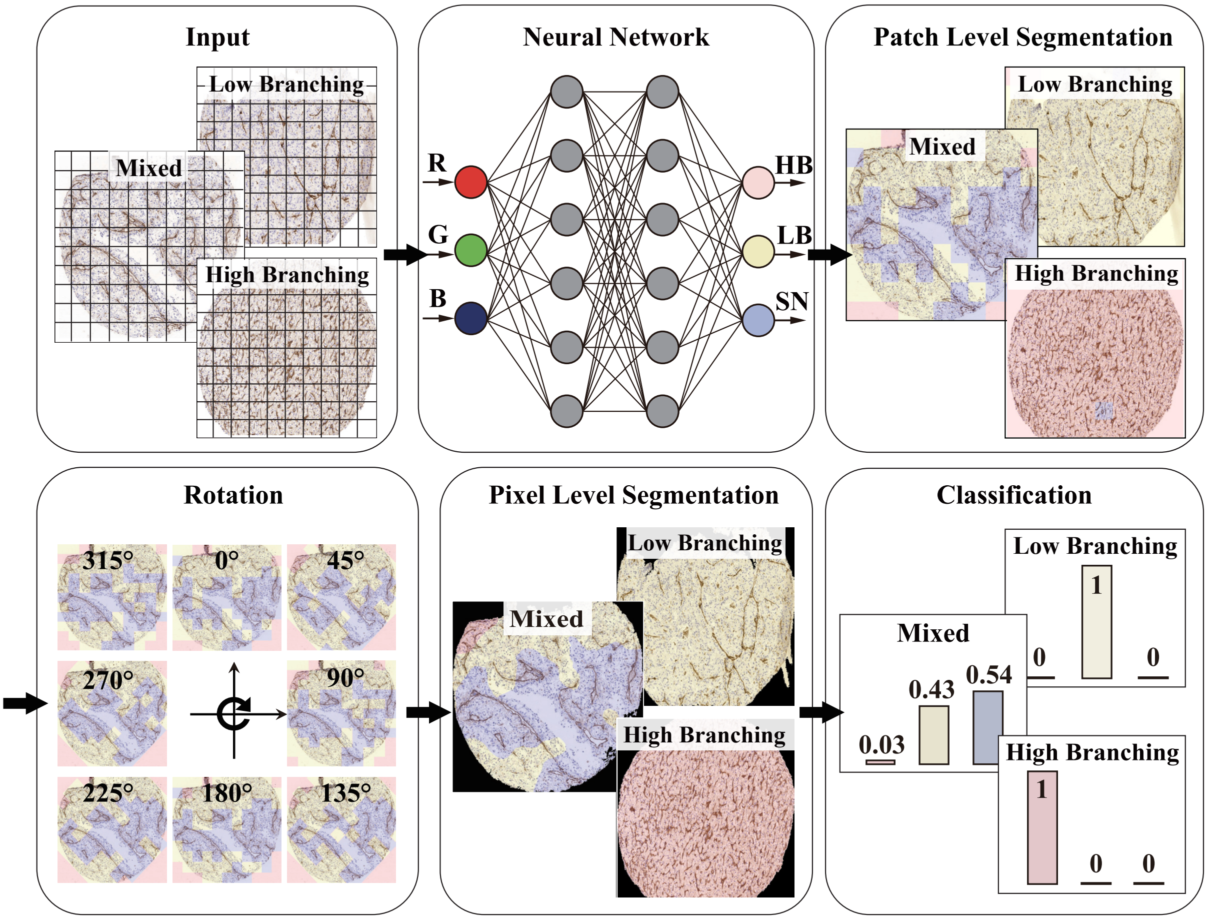


Figure 1: Detailed workflow of PropSegNet.

#### Experimental Setup

##### Dataset

The dataset used in this study was collected at the University Hospital Bonn. It contains 325 histopathology images of renal cell carcinoma (RCC), stained with CD31 and scanned at $10\times$ magnification. An experienced pathologist, with over 20 years of practice, annotated all slides. The images were classified into three vascular patterns: high branching (136 images), low branching (83 images), and sinusoid (63 images). Each image was analyzed individually. Its final label was determined by the most dominant pattern present. Additionally, 43 images were labeled as non-dominant. These showed a probability difference of less than 20% between at least two classes, making it difficult to assign a single dominant label. We used five-fold cross-validation in the experiments. In each fold, 80% of the data is used for training and 20% for testing.

##### Implementation Details

**Pre-processing.** To reduce visual variability and enhance model ignore non-tissue area, we implemented a pre-processing pipeline: stain normalization and background removal. In digital pathology, one of the major challenges lies in the variability of staining schemes, which introduces significant color and intensity inconsistencies in histopathology images. These variations stem from differences in slide preparation, staining batches, and scanner settings, and have been shown to negatively impact the performance of computer-aided diagnostic systems [1, 2]. To solve this problem, we used Vahadane's staining normalization technique [3], which improves the color consistency across samples. This approach employs color deconvolution to separate stain channels and applies sparse non-negative matrix factorization (NMF) to learn stain-specific features. Each input image is then normalized by aligning its NMF components with a target image, ensuring staining consistency across different histopathology slides.

Following normalization, we removed the background to eliminate large white margins and isolate tissue regions. Each image was converted to grayscale and binarized using inverse thresholding. The largest connected contour was extracted, and a bounding box was used to crop the image. This operation removes irrelevant background information and improves learning efficiency by focusing the model on tissue areas.

**Training Procedure.** Each slide was pre-processed to remove background regions and uniformly resized to a fixed resolution of $1500 \times 1500$ pixels. To enable fine-grained analysis of localized vascular patterns in high-resolution RCC slides, each image was subdivided into smaller patches. This patch-based formulation allows the model to learn detailed morphological features across spatial regions and generate dense, patch-level predictions that contribute to the final segmentation mask.

To capture fine-grained contextual information and reduce information loss at patch boundaries, overlapping patches were employed during training. Specifically, the image was divided into patches of size $224 \times 224$ pixels using a fixed stride of 150 pixels. This configuration resulted in a 37-pixel overlap on each side between adjacent patches. Zero-padding of 37 pixels was then applied to all sides of the image prior to patch extraction. This padding increased the overall image resolution to $1574 \times 1574$ pixels, enabling the generation of a regular $10 \times 10$ grid, resulting in 100 patches per slide.

During training, data augmentation was applied to enhance generalization and robustness. At the whole-slide level, vertical and horizontal flips were used, each with a probability of 0.5. At the patch level, random rotations were introduced with discrete angles of $0°$, $30°$, $60°$, $90°$, $120°$, or $150°$. In addition, Gaussian blur was applied using a kernel size of 11, with the standard deviation $\sigma$ randomly sampled from the interval $[0.05, 0.1]$. To simulate variations in imaging brightness, brightness jittering was employed with a multiplicative factor of 0.05. All input images were normalized using the mean and standard deviation values from the ImageNet dataset to maintain compatibility with the pre-trained backbone. The network was trained for 50 epochs using a batch size of 4.

**Post-processing.** Due to the small patch size of $224 \times 224$, the assignment of a pixel to a certain pattern is challenging. Moreover, there might be two or even three patterns with similar class probabilities, indicating that each patch may contain at least two predominant patterns. Therefore, delineating the locations of different classes within each patch is necessary. However, the image labels provided only a rough estimate of the cancer pattern, which further increased the uncertainty of the predictions. To improve the accuracy of the predictions, test time augmentation is employed to obtain the final pattern of each pixel. For this reason, each image is rotated with an angle $\{0°, 1°, \ldots, 359°\}$. After rotation, each slide was divided into 100 new patches of size $224 \times 224$, as depicted in Figure 1. Then, by aligning the predictions with the original image through reverse rotation, each pixel is assigned 360 labels representing the branching patterns. The final assignment of a pixel to a pattern is based on the majority of the votes, which ultimately results in a segmentation mask. Finally, the probability of each pattern is calculated based on the relative size of the segmentation masks.

#### Model Details

In this work, we propose a deep learning-based framework for segmenting and quantifying vascular branching patterns in RCC slides. Notably, our model is trained using only the global, slide-level estimates of pattern proportions, without any pixel-wise or patch-level annotations. The following subsections describe the model in detail.

##### Patch Weighting by Tissue Density

Histopathology images often contain large areas of background with minimal or no tissue content, especially after patch cropping. Treating all patches equally in this context can mislead the model, as background-dominated patches contribute little to the learning objective, yet are weighted the same as tissue-rich regions. To address this, we introduce a patch-wise weighting scheme based on the proportion of tissue pixels within each patch relative to the entire image. The weighting coefficient $\lambda_i$ for each patch $i$ is computed as:

$$\lambda_i = \frac{\text{number of tissue pixels in patch}_i}{\text{total number of tissue pixels}}.$$

Note that the patch size considered for the $\lambda_i$ calculation was $150 \times 150$ pixels, corresponding to the center region of each $224 \times 224$ patch. This size reflects the effective non-overlapping area determined by the stride of 150 pixels used during patch extraction.

##### Transformer Model

We employ the Swin Transformer architecture [4], pre-trained on ImageNet, as the backbone for both proportion estimation and segmentation tasks. The model utilizes a hierarchical design and localized self-attention mechanism to effectively capture both fine-grained and contextual representations in histopathology images.

The overall architecture is composed of four sequential stages, as illustrated in Figure 2. During the initial embedding step, each input image is partitioned into non-overlapping patches of size $4 \times 4$ pixels, resulting in a feature vector of dimensionality 48 ($4 \times 4 \times 3$). These features are then projected into a 96-dimensional space using a linear embedding layer, producing a compact yet informative representation that preserves local structural information.

Starting from stage 2, a *PatchMerging* layer is applied to aggregate features from adjacent $2 \times 2$ patches, effectively reducing spatial resolution while doubling the number of channels. This enables hierarchical abstraction and efficient representation learning across multiple scales.

Unlike standard transformer architectures that employ global self-attention and suffer from high computational overhead, Swin Transformer introduces a shifted window attention mechanism. In this approach, self-attention is first computed within non-overlapping $7 \times 7$ windows. To promote cross-window interactions, the window partitioning is shifted by half the window size (i.e., 3 pixels) in alternate layers. This design facilitates both local and global information exchange while maintaining linear complexity relative to image size. The combination of hierarchical feature extraction and the shifted windowing strategy allows the model to learn rich spatial dependencies at multiple scales, which is essential for modeling high-resolution pathology images.

For inference, the global class probabilities are computed by aggregating the outputs of the patch-level predictions across the full image:

$$(p^{HB}, p^{LB}, p^{SN}) = \sum_{i=1}^{N} \lambda_i \cdot \mathrm{Net}(\mathrm{patch_i}),$$

where $p^{HB}$, $p^{LB}$ and $p^{SN}$ denote the predicted probability on the level of the entire image for high branching, low branching, and sinusoid, respectively. Note that $\lambda_i$ is the proportion of tissue pixels in patch$_i$, and Net(patch$_i$) represents the output of the network for this patch.

As a loss function, we incorporated the mean squared error (MSE) between the predicted probabilities ($p^{HB}, p^{LB}, p^{SN}$) and image label ($P^{HB}, P^{LB}, P^{SN}$) as the objective function. The model was trained for 50 epochs with a batch size of 2 and a learning rate of $3 \cdot 10^{-5}$ using the AdamW optimizer [5]. During training, the initialization learning rate was decreased by using the cosine-annealing learning rate scheduler [6].

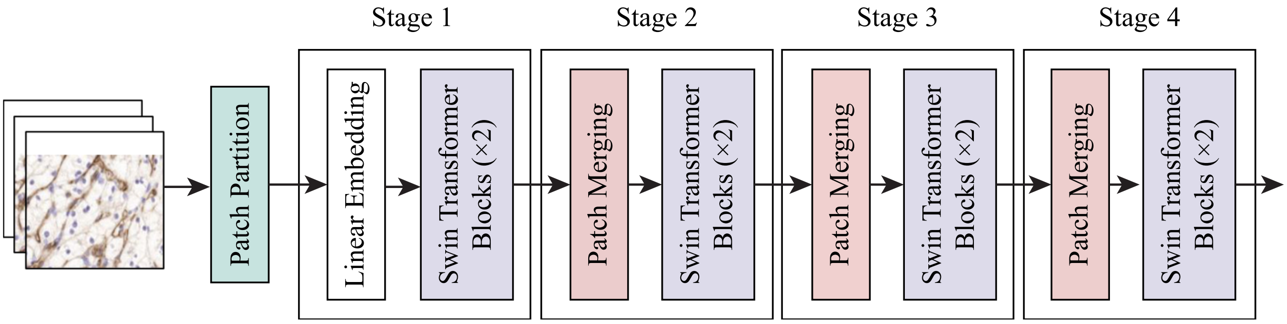


(a)General architecture of the swin Transformer

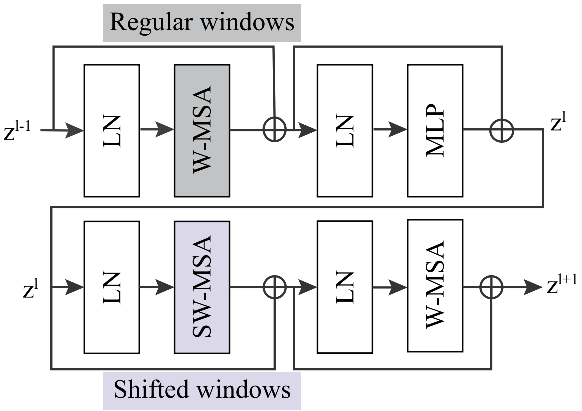


(b)Two Successive Swin Transformer Blocks

Figure 2: Architecture of the Swin Transformer. (a) General architecture of the Swin Transformer; (b) Two successive Swin Transformer Blocks (Swin Transformer Block with alternating regular and sliding windows).

#### Experimental Evaluation

##### Evaluation Metrics

For a particular image slide of the dataset, the global probability across the entire image is defined as the proportion of each pattern within the entire tissue. The average results of the five-fold training models were rigorously evaluated by an experienced pathologist to benchmark the model's performance. The evaluation metrics used in this paper are accuracy, precision, recall, and F1-score. Note that TP refers to true positive, i.e., the predominant prediction is correctly classified as belonging to the positive class. The other metrics are defined analogously: true negative (TN: correct negative class identification), false positive (FP: incorrect positive class prediction), and false negative (FN: incorrect negative class prediction). The performance metrics used throughout this manuscript are listed in Table 1.

| Metrics | Definition |
|---|---|
| Accuracy | $ACC = \frac{\sum_{i=1}^{N} \mathrm{TP}_i}{N}$ |
| Precision | $Pr = \frac{\mathrm{TP}}{\mathrm{TP+FP}}$ |
| Recall | $R = \frac{\mathrm{TP}}{\mathrm{TP+FN}}$ |
| F1-score | $F1 = 2 \cdot \frac{Pr \cdot R}{Pr + R}$ |

Table 1: Evaluation metrics

##### Results

The metrics presented in Table 2 were computed by the five-fold training models. This table reports the accuracy, precision, recall, and F1-score for each fold, as well as their averages. The average values provide a comprehensive evaluation of the classification performance, ensuring the robustness of the model.

| Metrics | Fold-1 | Fold-2 | Fold-3 | Fold-4 | Fold-5 | Average |
|---|---|---|---|---|---|---|
| Accuracy | 92.73% | 96.36% | 93.36% | 94.44% | 90.91% | 94.16% |
| Precision | 93.45% | 96.34% | 96.34% | 93.469% | 91.97% | 94.36% |
| Recall | 91.02% | 96.08% | 96.08% | 94.56% | 88.94% | 93.34% |
| F1-score | 91.51% | 96.03% | 96.03% | 93.98% | 89.85% | 93.48% |

Table 2: Average results of the five-fold training models

The confusion matrix in Figure 3 summarizes the model's classification performance across the three vascular patterns: high branching (HB), low branching (LB), and sinusoid (SN). The results correspond to the average confusion matrix computed over the five folds of the cross-validation. Each element in the matrix represents the percentage of slides where the true pattern matches the prediction. The matrix shows that the model behaves well in correctly classification, with a high percentage of TP on the diagonal.

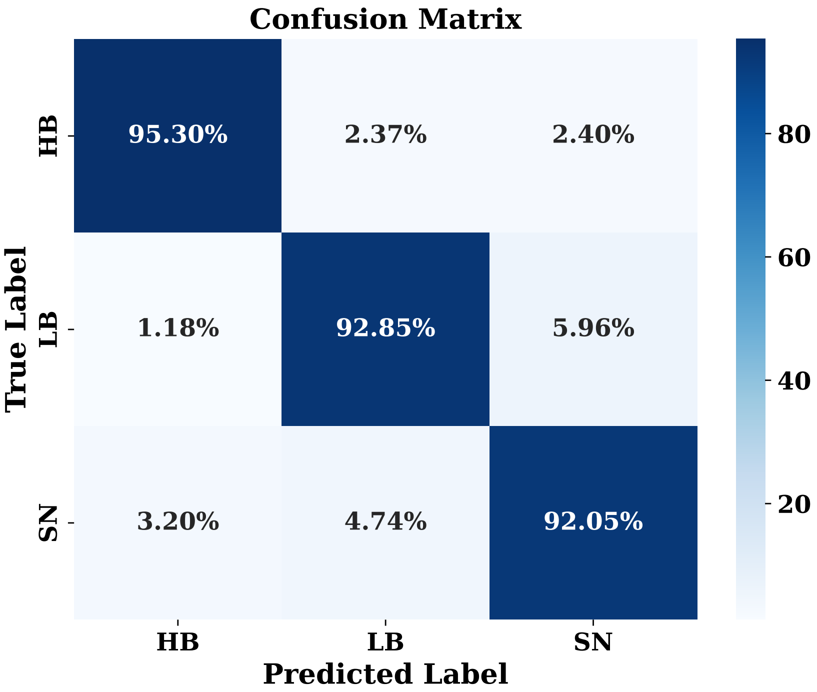


Figure 3: Average confusion matrix
