## Supplementary_Table_1_PropSegCohort for "Low-branching vessel architecture shapes immune cell niches and predicts immune responses in renal cancer"

**Supplementary Table 1.** Basic patient characteristics of ccRCC cohorts used for the development of PropSegNet.

| Patient characteristics | PropSegNet cohort A  (n= 354) | | PropSegNet cohort B  (n=265) | |
| --- | --- | --- | --- | --- |
| Age |  |  |  |  |
| Median (years), range (years) | 66 | (13 - 89) |  | (30-86) |
| Sex |  |  |  |  |
| Male, n (%) | 234 | (66.1) | 168 | (63.4) |
| Female, n (%) | 120 | (33.9) | 94 | (35.5) |
| NA |  |  | 3 | (1.1) |
| Grading, n (%) |  |  |  |  |
| 1 | 48 | (13.6) | 49 | (18.5) |
| 2 | 240 | (67.8) | 145 | (54.7) |
| 3 | 52 | (14.7) | 49 | (18.5) |
| 4 | 12 | (3.4) | 21 | (7.9) |
| NA | 2 | (0.6) | 1 | (0.4) |
| Tumor staging, n (%) |  |  |  |  |
| T1 | 222 | (62.7) | 153 | (57.7) |
| T2 | 25 | (7.1) | 19 | (7.2) |
| T3 | 102 | (28.8) | 89 | (33.6) |
| T4 | 5 | (1.4) | 3 | (1.1) |
| NA | 0 | (0) | 1 | (0.4) |
| Lymph node, n (%) |  |  |  |  |
| 0 | 224 | (63.3) |  |  |
| 1 | 17 | (4.8) |  |  |
| 2 | 80 | (22.6) |  |  |
| NA | 33 | (9.3) |  |  |
| Distant metastases, n (%) |  |  |  |  |
| Yes | 34 | (9.6) |  |  |
| No | 279 | (78.8) |  |  |
| NA | 41 | (11.6) |  |  |

NA: not available.
