## Supplementary_Table_6_ALI_cohort_pat_characteristics for "Low-branching vessel architecture shapes immune cell niches and predicts immune responses in renal cancer"

**Supplementary Table 6.** Basic patient characteristics of ccRCC cohort used for air-liquid-interface (ALI) cultures of patient-derived tumor fragments (PDTF).

|  | ALI PDTF ccRCC cohort  (*n*=30) | |
| --- | --- | --- |
| Age | 66 | (39-83) |
| Median (years), range (years) |  |  |
| Sex |  |  |
| Male, *n* (%) | 19 | (63.3) |
| Female, *n*(%) | 11 | (36.7) |
| NA |  |  |
| Grading, *n* (%) |  |  |
| 1 | 7 | (23.3) |
| 2 | 13 | (43.3) |
| 3 | 7 | (23.3) |
| 4 | 3 | (10) |
| NA |  |  |
| Tumor staging, *n* (%) |  |  |
| T1 | 21 | (70) |
| T2 | 2 | (6.66) |
| T3 | 7 | (23.3) |
| T4 | 0 | (0) |
| NA |  |  |
| Lymph node, *n* (%) |  |  |
| 0 | 29 | (96.7) |
| 1 | 1 | (3.3) |
| 2 | 0 | (0) |
| NA |  |  |
| Distant metastases, *n* (%) |  |  |
| Yes | 1 | (3.3) |
| No | 29 | (96.7) |
| NA |  |  |

NA: not available.
