## Supplementary_Table_8_QiagSeq for "Low-branching vessel architecture shapes immune cell niches and predicts immune responses in renal cancer"

**Supplementary Table 8.** QIAseq targeted DNA custom gene panel: targeted regions

| **Gene** | **refseq transcript ID**  **RefSeq** | **Ensembl transcript ID**  **Ensembl** | **Number of targeted exons** | **Main hotspot codons** |
| --- | --- | --- | --- | --- |
| *ARID1A* | NM_006015 | ENST00000324856 | 20 | whole coding sequence |
| *BAP1* | NM_004656 | ENST00000460680 | 17 | whole coding sequence |
| *KDM5C* | NM_004187 | ENST00000375401 | 26 | whole coding sequence |
| *MTOR* | NM_004958 | ENST00000361445 | 58 | whole coding sequence |
| *PTEN* | NM_000314 | ENST00000371953 | 9 | whole coding sequence |
| *PBRM1* | NM_018313 | ENST00000394830 | 30 | whole coding sequence |
| *SETD2* | NM_014159 | ENST00000409792 | 21 | whole coding sequence |
| *TP53* | NM_001276761 | ENST00000269305 | 11 | whole coding sequence |
| *VHL* | NM_000551 | ENST00000256474 | 3 | whole coding sequence |
