## Supplementary_Table_2_3_advanced_RCC_characteristics for "Low-branching vessel architecture shapes immune cell niches and predicts immune responses in renal cancer"

**Supplementary Table 2_3.** Basic patient characteristics of advanced stage ccRCC cohort used for PopSeqNet validation.

61 patients (67 samples: 48 primary tumors, 6 paired distant metastases, 13 unpaired distant metastases).

|  | Advanced stage ccRCC cohort  (n=61) | |
| --- | --- | --- |
| Age |  |  |
| Median (years), range (years) | 65 | (48 - 79) |
| Sex |  |  |
| Male, *n* (%) | 23 | (37.7) |
| Female, *n* (%) | 11 | (18.0) |
| NA (%) | 27 | (44.3) |
| Grading, *n* (%) |  |  |
| 1 | 0 | 0 |
| 2 | 13 | (21.3) |
| 3 | 26 | (42.6) |
| 4 | 11 | (18.0) |
| NA | 11 | (18.0) |
| Tumor staging, *n* (%) |  |  |
| T1 | 15 | (24.6) |
| T2 | 9 | (14.7) |
| T3 | 23 | (37.7) |
| T4 | 2 | (3.3) |
| NA | 12 | (19.7) |
| Lymph node, *n* (%) |  |  |
| 0 | 32 | (52.4) |
| 1 | 5 | (8.2) |
| 2 | 12 | (19.7) |
| NA | 12 | (19.7) |
| Distant metastases, *n* (%) |  |  |
| Yes | 61 | (100) |
| No | 0 | 0 |
| NA |  |  |
| Therapy; *n* (%) |  |  |
| 1 | 27 | (44.3) |
| 2 | 15 | (24.6) |
| 3 | 1 | (1.6) |
| 4 | 6 | (9.8) |
| NA | 12 | (19.7) |
| Response (Outcome Staging 1) |  |  |
| PD | 22 | (36.1) |
| PR | 9 | (14.8) |
| SD | 10 | (16.4) |
| MR | 3 | (4.9) |
| CR | 1 | (1.6) |
| NA | 16 | (26.2) |

Therapy: 1= nivolumab; 2 = nivolumab + ipilimumab; 3 = pembrolizumab; 4 = pembrolizumab + axitinib;

Response: PD = progressive disease, PR = partial response, SD = stable disease, MR = mixed response, CR = complete response. NA: not available.
